# Potency and timing of antiviral therapy as determinants of duration of SARS CoV-2 shedding and intensity of inflammatory response

**DOI:** 10.1101/2020.04.10.20061325

**Authors:** Ashish Goyal, Cardozo-Ojeda, E. Fabian, Joshua T. Schiffer

## Abstract

Treatments are desperately needed to lower the hospitalization and case fatality rates of SARS CoV-2 infection. In order to meaningfully impact the COVID-19 pandemic, promising antiviral therapies must be identified within the next several months. However, the number of clinical trials that can be performed in this timeframe is limited. We therefore developed a mathematical model which allows projection of all possible therapeutic approaches. Our model recapitulates off-treatment viral dynamics and predicts a three-phase immune response. Addition of treatment with remdesivir, hydroxychloroquine, neutralizing antibodies or cellular immunotherapy demonstrates that if *in vivo* drug potency is high, then rapid elimination of virus is possible. Potent therapies dosed soon after peak viral load when infected people typically develop symptoms, are predicted to decrease shedding duration and intensity of the effector immune response, but to have little effect on viral area under the curve, which is driven by high levels of early SARS CoV-2 replication. Potent therapy dosed prior to peak viral load, when infection is usually pre-symptomatic, is predicted to be the only option to lower viral area under the curve. We also identify that clinically meaningful drug resistance is less likely to emerge with a highly potent agent that is dosed after peak viral load. Our results support an early test and treat approach for COVID-19, but also demonstrate the need to identify early viral shedding kinetic features that are the most predictive surrogates of clinical severity and transmission risk.

**One Sentence Summary:** We developed a mathematical model to predict the outcomes of different possible COVID-19 treatments.

## Introduction

The COVID-19 pandemic is a devastating historical event which is currently impacting nearly all of mankind. SARS CoV-2 incidence is surging in numerous cities and countries across the globe (*1*), and infection carries a high mortality rate, particularly among the elderly (*2-4*).

While social distancing has slowed and even eliminated many local epidemics, it is not an economically viable long-term strategy (*5*). There is no evidence of widespread herd immunity and a vaccine is unlikely to be developed and implemented within the next 18 months.

Therefore, second and third waves of infection are likely to occur over the next two years (*6*).

It is imperative that optimal treatment strategies of COVID-19 are identified during the first wave of infection to ensure that the case fatality rate is lower during subsequent local epidemics. To date, selection of antiviral agents has been empirical and guided by limited or absent data. The window to conduct definitive clinical trials is likely to be narrow given that flatten the curve efforts are predicted to slow local epidemics, albeit temporarily. Therefore, effective tools are urgently needed to optimize the design of these trials.

Here we use mathematical models to project the possible impact of two small molecular agents, remdesivir and hydroxychloroquine; as well as broadly neutralizing antibodies and cellular immunotherapies. The goal of our models is to interpret emerging clinical trial data, and in turn to perfect subsequent trials in terms of selection of antiviral agents, timing of therapy, dosage, treatment duration, avoidance of drug resistance and selection of virologic endpoints.

Overall, our simulations support initiation of therapy soon after symptoms develop and also suggest the urgent need for studies to identify virologic surrogates of SARS CoV-2 severity.

## Results

### SARS CoV-2 natural history

We used four datasets of SARS CoV-2 shedding in the absence of effective treatment to develop and validate a mathematical model. This data included 25 infected people: 11 from Singapore (*7*), 9 from Germany (*8*), one from Korea (*9*), and 4 from France (*10*) **(Fig 1)**.

**Figure 1.**
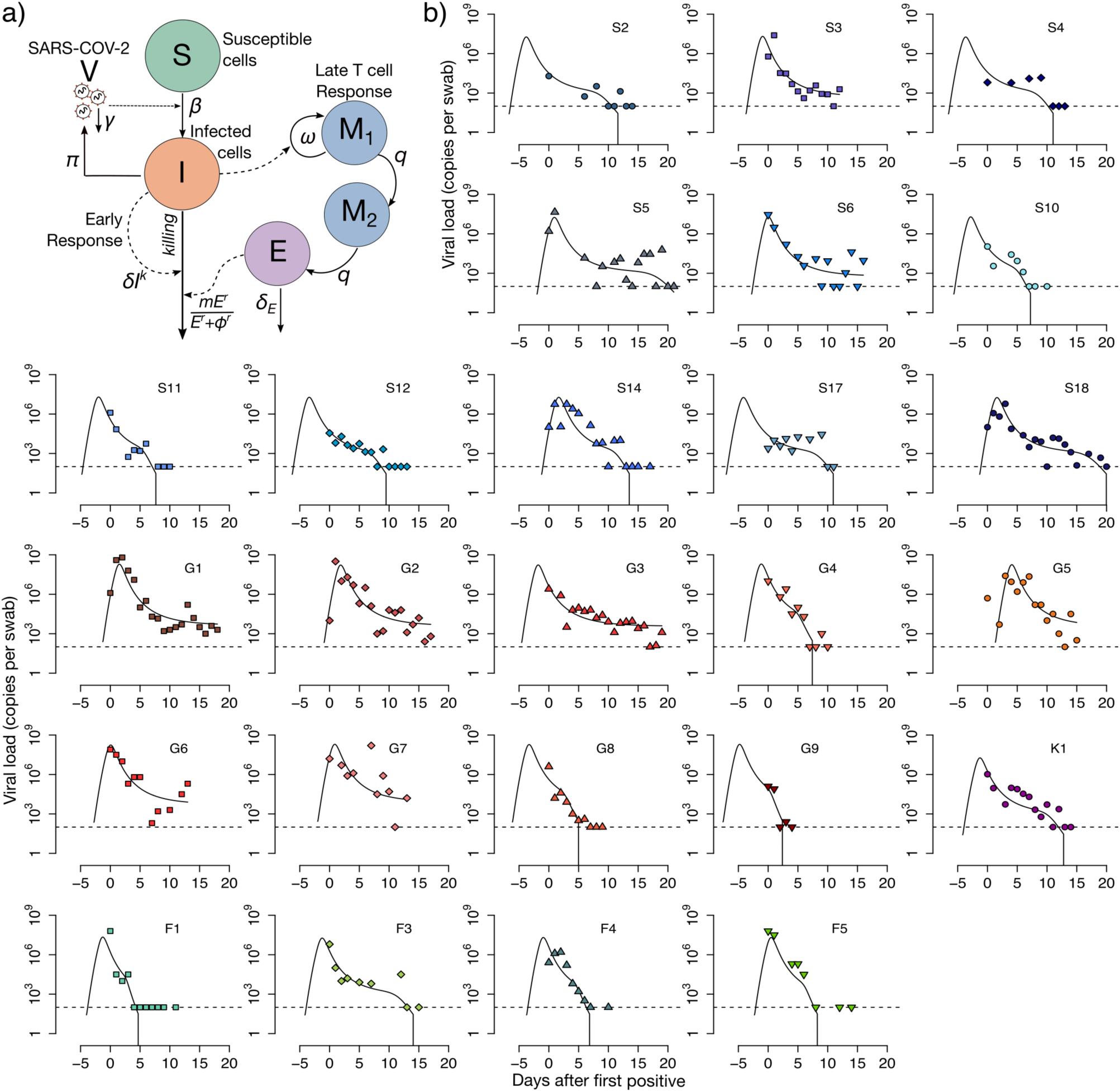
Mathematical model recapitulation of untreated SARS CoV-2 kinetics. **A**. Mathematical models schematic including infection of susceptible cells (S), production of virus by (V) by infected cells (I), an early density-dependent immune response governed by exponent k, and a mounting T cell response with sequential populations of T cells (M1, M2, M3 and E) which kill infected cells when above a certain threshold. **B**. Model fit to individual data. Shapes are individual viral loads and lines are model load projections. S = Singapore; G = Germany; K = Korea; F = France.

Notably, sampling techniques differed across studies. In Singapore, Korea and France, samples were obtained with nasopharyngeal swabs, whereas in Germany viral loads were measured directly from sputum. Shedding was notable for an early peak, followed by three phases of viral decay including a rapid initial decline from peak, a slower period of decay of variable length, followed by a third abrupt elimination of the low levels of remaining virus. Of note, we only captured the viral peak as well as the final rapid clearance phase in a subset of study participants. There was one example of substantial, transient viral re-expansion (G7 in **Fig 1**).

### SARS CoV-2 mathematical model

We developed a series of ordinary differential equations to fit to the viral load data **(Fig. 1a, Methods and Materials)**. The equations capture the coupled interactions of susceptible cells, infected cells, SARS CoV-2 and a mounting immune response. In keeping with the standard viral dynamics model (*11, 12*), virus enters susceptible cells and converts them to infected dells which then produce virus at a fixed rate. Based on model fitting, we included two immune responses. The first accounts for the rate of infected cell elimination by the innate immune system and is governed by an exponent; in keeping with prior research, we refer to this as the density-dependent immune response (*13, 14*). The second phase is a slower cytolytic response in which per cell killing rate saturates once the total number of effector cells exceeds a certain level. We model this with stages of presumed effector cell precursors which differentiate at rate *q* as a method to calibrate timing.

Our model reproduced viral load kinetics in all 25 participants **(Fig. 1b)**. In certain cases, the model only fit to available data from the later stages of shedding, whereas it recapitulated the entirety of viral expansion, peak and decelerating clearance for several study participants (S5, S14, S18, G1, G2, G5, G7). In keeping with observations from a recent clinical trial (*15*), low level shedding continued past 20 days for some German participants (G1, G2, G3, G5, G6), whereas viral elimination occurred in the remaining infected people.

### Timing of innate and acquired cytolytic responses

We continuously quantified the value of the immune terms in all 25 participants: the per cell killing rate and total number of cells killed per day was extremely high during the first several days of infection in all participants **(Fig. 2a, b)**, coinciding with peak viral load. The cytolytic immune response initiated at various timepoints across participants (day 5-17) and led to lower per cell and total killing rates relative to the innate response but was sufficient to eliminate remaining infection **(Fig. 2b)**.

**Figure 2.**
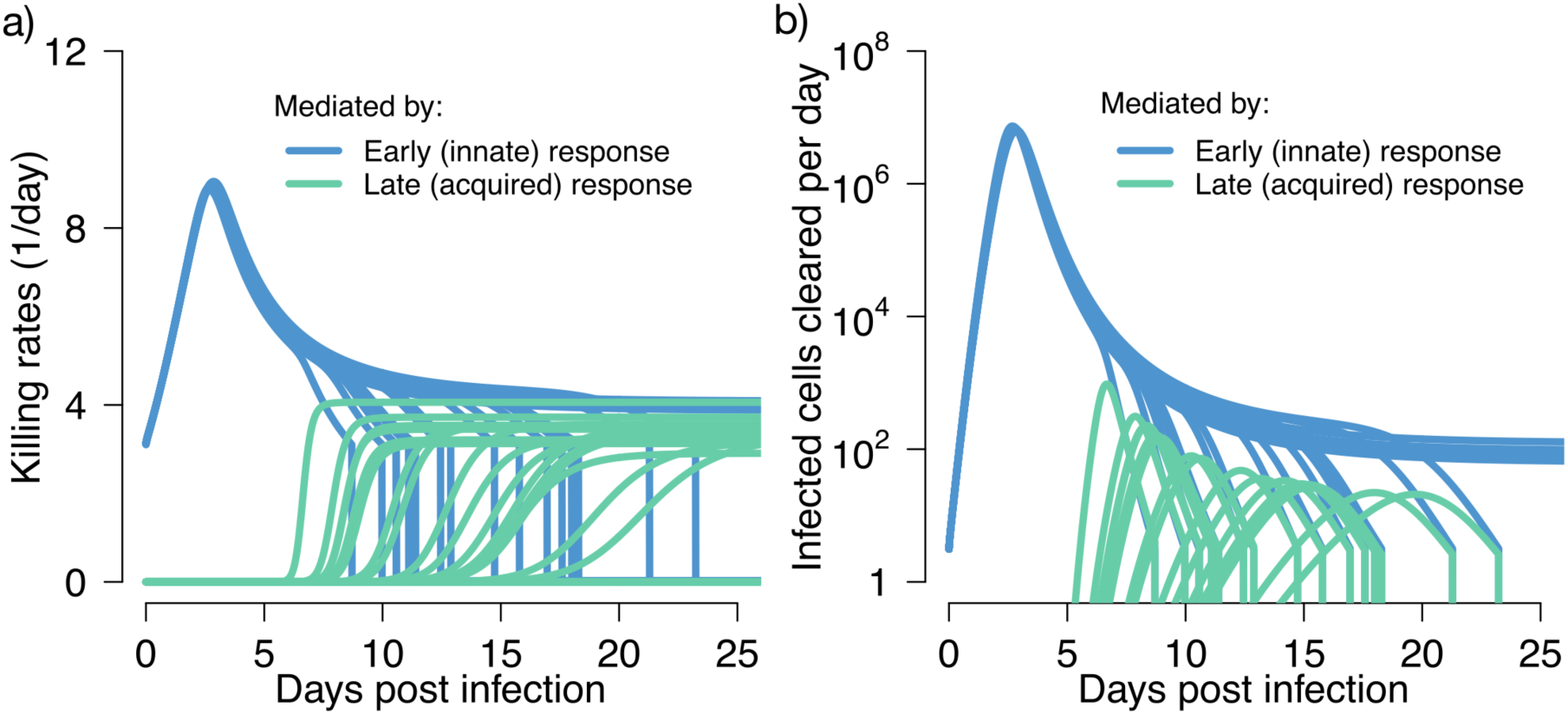
Early innate and late acquired killing rates of SARS CoV-2 infected cells. Model projections of rates in 15 participants who cleared viral shedding. **A**. Per cell death rate mediated by innate responses (blue) and acquired responses (green). **B**. Total death rate mediated by innate responses (blue) and acquired responses (green).

### Remdesivir pharmacokinetics and pharmacodynamics

We developed a pharmacokinetic / pharmacodynamic model of remdesivir **(Fig. 3a)**, a broad-spectrum nucleotide analogue that targets SARS CoV-2 replication in infected cells (*16*). The model links intravenous administration with plasma levels of free drug and concentrations of the drug’s active nucleotide-triphosphate component (NTP) observed within PBMCs in non-human primates (*17*), and captures the slow decay of NTP within this compartment **(Fig. 3b)**. With multiple doses, we project stable levels of NTP in target cells over time **(Fig. 3c)** followed by slow decay after cessation of treatment.

**Figure 3.**
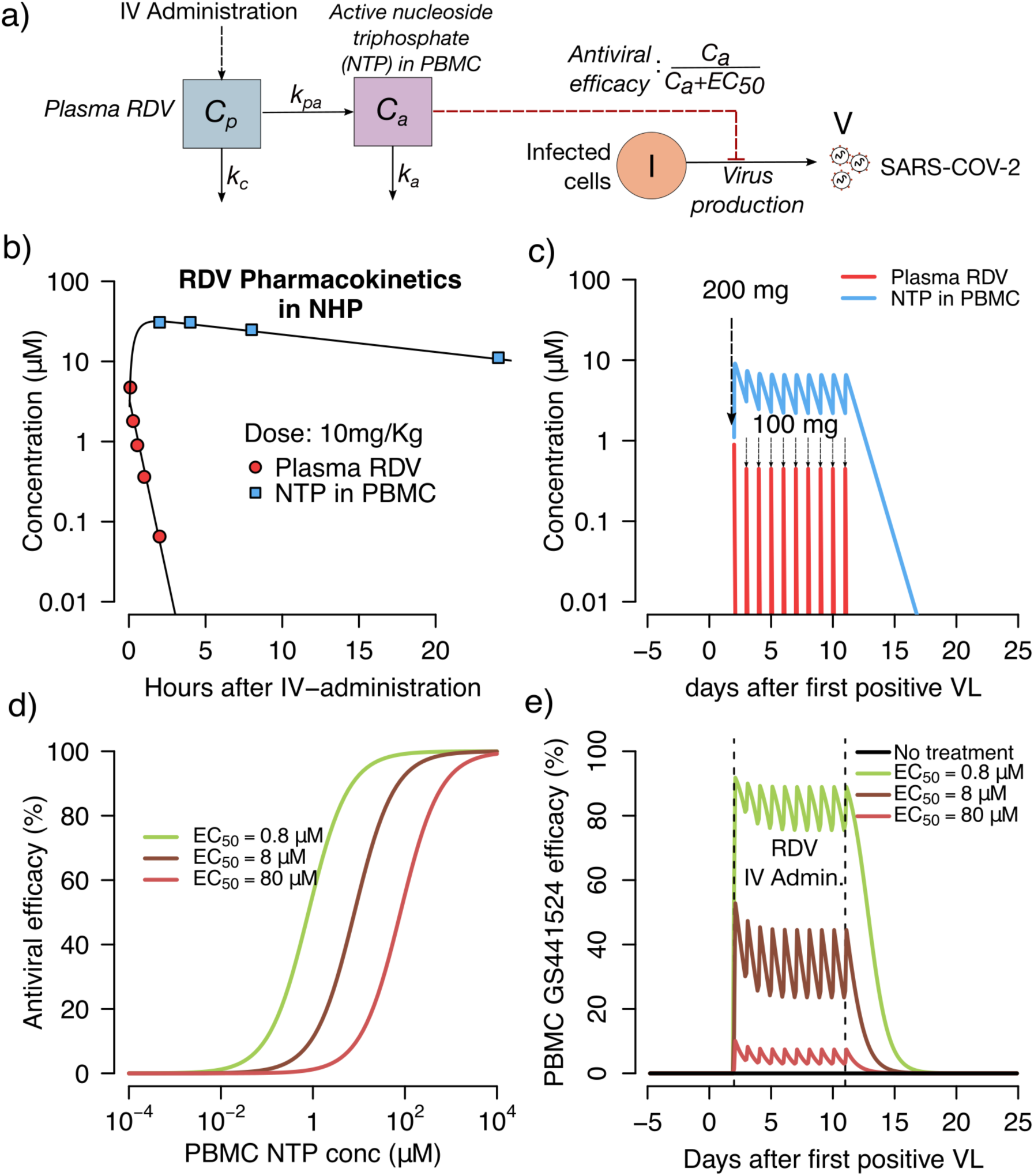
Projected pharmacokinetics and pharmacodynamics of remdesivir therapy. **A**. Complete model of remdesivir (RDV) including plasma levels of parent drug, intracellular levels of the active component (NTP) and antiviral efficacy of drug according to concentration. **B**. Projections of plasma RDV levels and intracellular NTP in PBMCs. Datapoints from non-human primate experiments are dots while lines are model projections. **C**. Simulated concentrations of the parent compound and intracellular levels of the active compound with a loading dose of 200 mg IV followed by 9 daily doses of 100 mg IV. **D**. Pharmacodynamic projections of antiviral efficacy according to drug concentration assuming different values for the *in vivo* EC50 of the drug. **E**. Combination simulations of pharmacokinetic and pharmacodynamic models demonstrating prolonged antiviral activity after dosing is stopped.

We next simulated possible dose response curves of antiviral efficacy, which capture the percentage of viral replication eliminated, according to micromolar concentration of drug. The percent of viral replication suppression at a given intracellular drug concentration is dependent on the intracellular EC50 of the drug, or the concentration of drug required to lower viral replication by 50% **(Fig 3d)**. Of note the intracellular EC50 is unknown for remdesivir, particularly *in vivo*, making predictions of clinical trial outcomes impossible.

Finally, we combined the PK and PD models as in **Fig. 3a** to project the percentage of viral replication inhibited over time at different assumed intracellular EC50 values (*18, 19*). With high assumed drug potency (EC50=0.8 uM), antiviral effect are sustained over the 10-day dosing interval and are maintained for a long period once drug delivery has stopped **(Fig. 3e)**. At higher assumed EC50 values, remdesivir potency is projected to be lower.

### Projections of SARS CoV-2 outcomes assuming remdesivir treatment during early and late symptomatic phases

We next simulated therapy at day 10 of infection **(Fig. 4a)**, when severely infected people often seek hospital care, and at day 5 of infection immediately after viral peak **(Fig. 4b)**, when infected people often become symptomatic. In both cases, when remdesivir *in vivo* potency was assumed to be high (EC50=0.8 uM), viral elimination occurred rapidly after initiation of therapy. This effect occurred because of unopposed removal of approximately 100-1000 infected cells per day by an ongoing innate immune response **(Fig. 2b)**.

**Figure 4.**
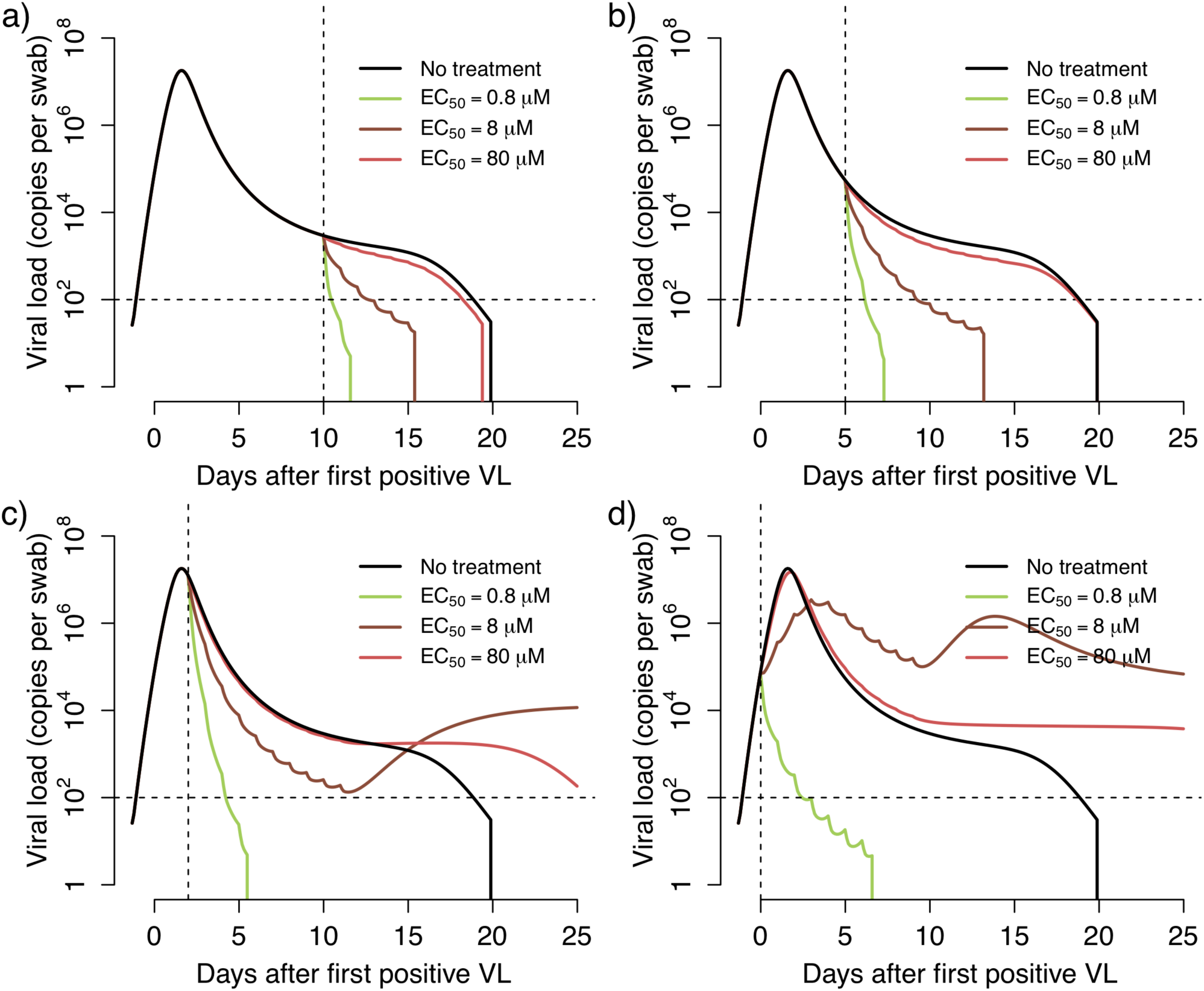
Treatment projections of a 10-day remdesivir course assuming different potency and timing of treatment. Each set of simulations is performed under assumptions of high, medium and low potency (EC50=0.8, 8 and 80 uM respectively). Treatment initiation at timepoints generally consistent with **A**. hospitalization (day 10 after first positive sample), **B**. first symptoms (day 5 after first positive sample), **C**. pre-symptomatic post-peak phase (day 2 after first positive sample) and **D**. pre-symptomatic pre-peak phase (day 0). Overall, early potent treatment limits duration of infection.

Simulations with the assumption of a less potent version of remdesivir (EC50= 8 or 80 uM) resulted in a lower viral clearance slope. This result implies that viral clearance slope in clinical trials can be used in concert with our model to directly estimate the *in vivo* intracellular EC50 value of remdesivir in each treated patient. The model can then be used to project the effect of increasing or decreasing doses in subsequent trials.

### Projections of SARS CoV-2 outcomes assuming extremely early remdesivir treatment during the pre-symptomatic phase

We next performed simulations of therapy at very early timepoints during infection at and prior to peak viral load. Most infected people are pre-symptomatic at this stage so these model realizations may most closely reflect implementation of a post exposure prophylaxis strategy in which some people are already in the very early stages of infection at the time of first dose. Under this scenario, extremely potent (EC50=0.8 uM) therapies at day 2 and 0 of infection resulted in immediate viral suppression **(Fig. 4c, d)**. With early, low or moderate potency treatment, the model predicted therapeutic failure with persistent SARS CoV-2 shedding due to insufficient early immunity against the virus **(Fig. 4d)**.

### Projections of short course remdesivir

We next repeated the above exercise with a shorter 5-day course of treatment. Results were similar though simulations under the assumption of extremely early initiation of therapy did not lead to full SARS CoV-2 suppression within this timeframe **(Fig. S1)**.

### Predictors of therapeutic efficacy for remdesivir

We next assessed which unknown variables in our therapeutic model were most predictive of relevant therapeutic outcomes. As independent variables, we selected *in vivo* intracellular EC50, because the potency of remdesivir against SARS CoV-2 in humans is unknown, and infection duration at the time of treatment initiation. As dependent variables, we selected shedding duration and viral area under the curve because it is unclear which of these outcomes is a stronger predictor of progression to cytokine storm and respiratory failure, as well as transmissibility, in infected people. We also included the final tally of effector cells as this outcome may also be predictive of likelihood of cytokine storm (*20*).

Early initiation of a highly potent therapy was predictive of lower shedding duration whether given in the pre-peak asymptomatic phase or in the post-peak symptomatic phase beyond day 2-4 of infection. However, extremely early initiation of a lower potency therapy was predicted to prolong shedding relative to no treatment **(Fig. 5a)**.

**Figure 5.**
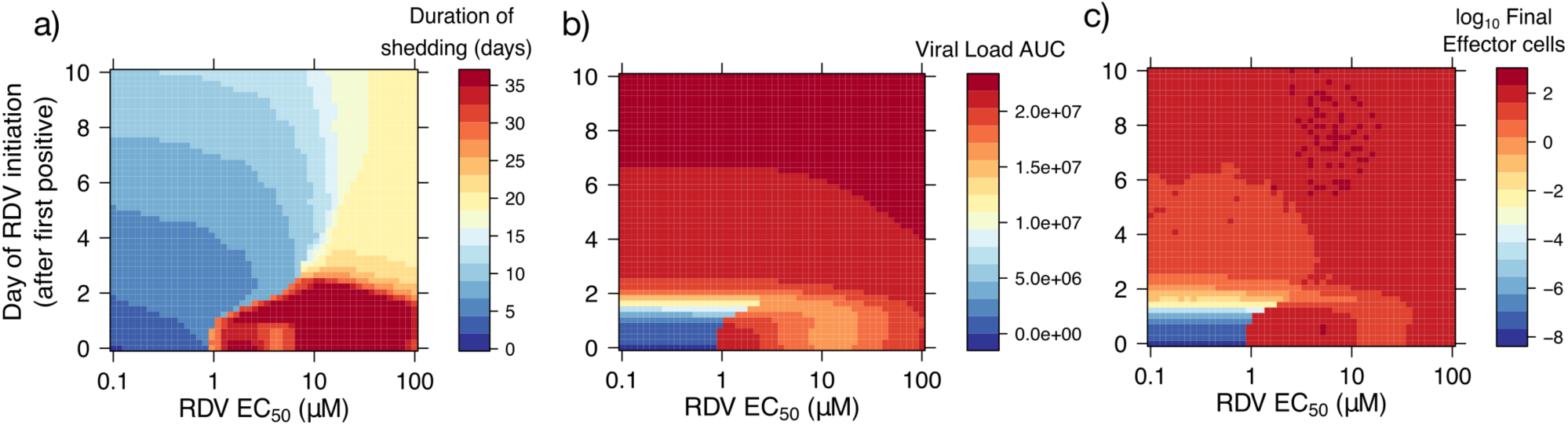
Predictors of SARS CoV-2 treatment outcomes with remdesivir. Heatmaps comparing variance in drug potency measured by *in vivo* EC50 (x-axis) and timing of treatment initiation (y-axis) for **A**. Shedding duration, **B**. viral load area under the curve (AUC) and **C**. extent of T cell response required for viral elimination. Potent therapy within the first 5 days of infection limits shedding duration and the extent of the T cell response. However, only extremely early therapy during the pre-symptomatic phase of infection lowers viral AUC. Sub-potent therapy given during the extremely early pre-symptomatic stage may extend shedding duration at lower viral loads by limiting the effector cell response.

Both high drug potency and extremely early treatment initiation during the pre-symptomatic stage of infection were required to significantly lower viral area under the curve. Even highly potent therapy during the earliest symptomatic phase at days 4-5 had only a slight impact on viral area under the curve, reflecting the fact that most virus and infected cells are generated during the first 2-3 days of SARS CoV-2 infection **(Fig. 5b)**.

Finally, initiation of a highly potent therapy within 6 days of infection lowered the extent of the effector cell response in our simulations **(Fig. 5c)** which may suggest that an early test and treat strategy could lower deleterious infection associated inflammation.

### Theoretical kinetics of drug resistant variants

Based on the mutation rate of positive ss RNA viruses of approximately 10^−5^ mutations per base pair per cell infection (*21*), and on the fact that two separate mutations may induce partial remdesivir resistance in SARS CoV-1 which in turn leads to a less fit virus (*22*), we estimated the probability that a drug resistant mutant would emerge during therapy. When we assumed a potent therapy (EC50=0.8), the model projects that while single and double mutants will emerge, they are unlikely to predominate or meaningfully extend duration of shedding if dosed during the symptomatic phase of disease, though resistance may emerge with early dosing **(Fig. 6a,b)**. However, if a moderate potency is assumed, then a single mutant with resistant is predicted to persist, particularly if therapy is initiated before or during viral peak **(Fig. 7a,b)**.

**Figure 6.**
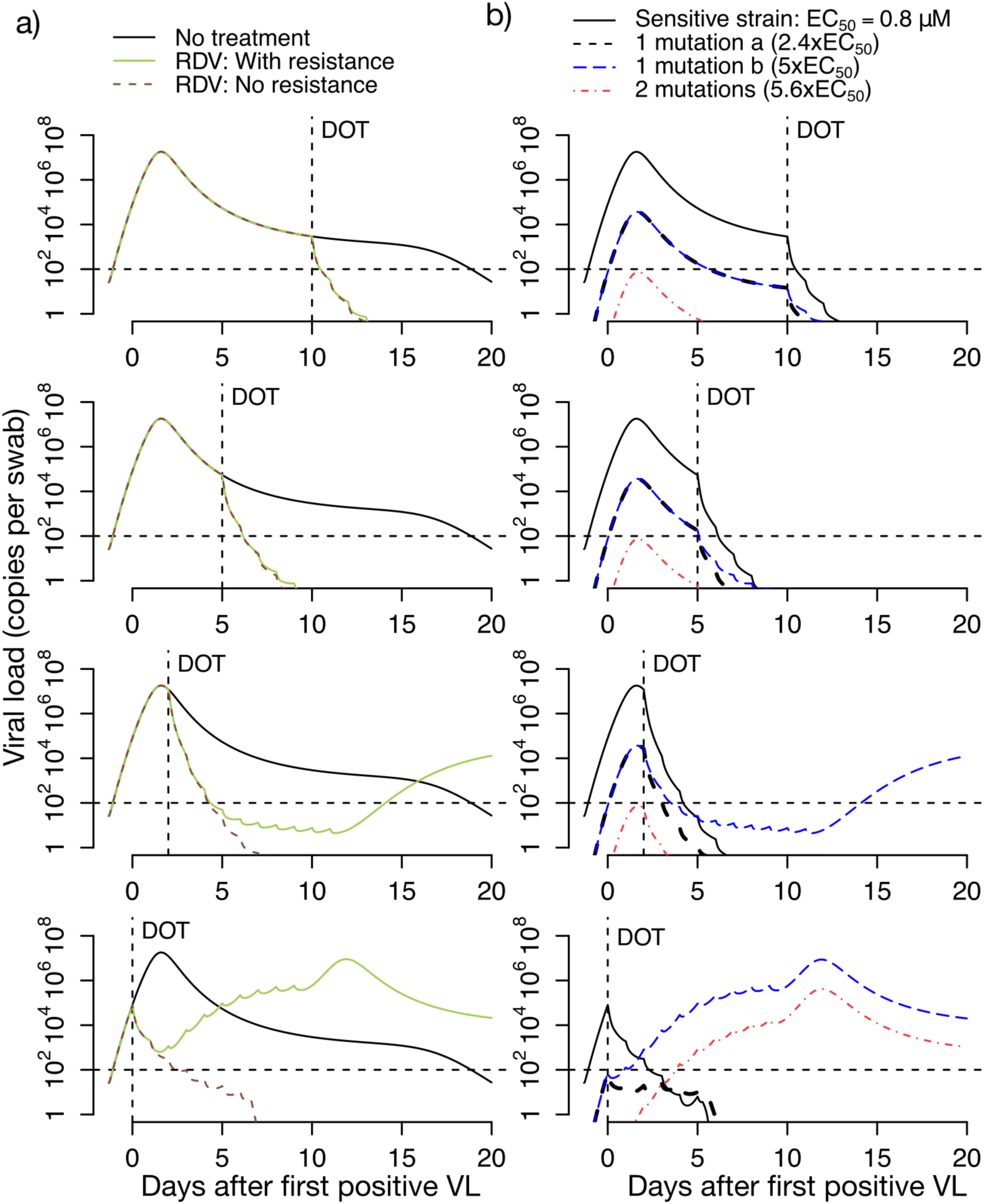
Projections of remdesivir drug resistance during therapy. Simulations are with high potency (EC50=0.8 uM) and the assumption that mutants confer partial drug resistance. Treatment initiation is at timepoints generally consistent with hospitalization (day 10 after first positive sample), first symptoms (day 5 after first positive sample), pre-symptomatic post-peak phase (day 2 after first positive sample) or pre-symptomatic pre-peak phase (day 0). **A**. Projections of no treatment, treatment with no assumed drug resistance, and treatment with assumed drug resistance. **B**. Projections of assumed drug resistance with trajectories of sensitive strains, single mutants and double mutants.

**Figure 7.**
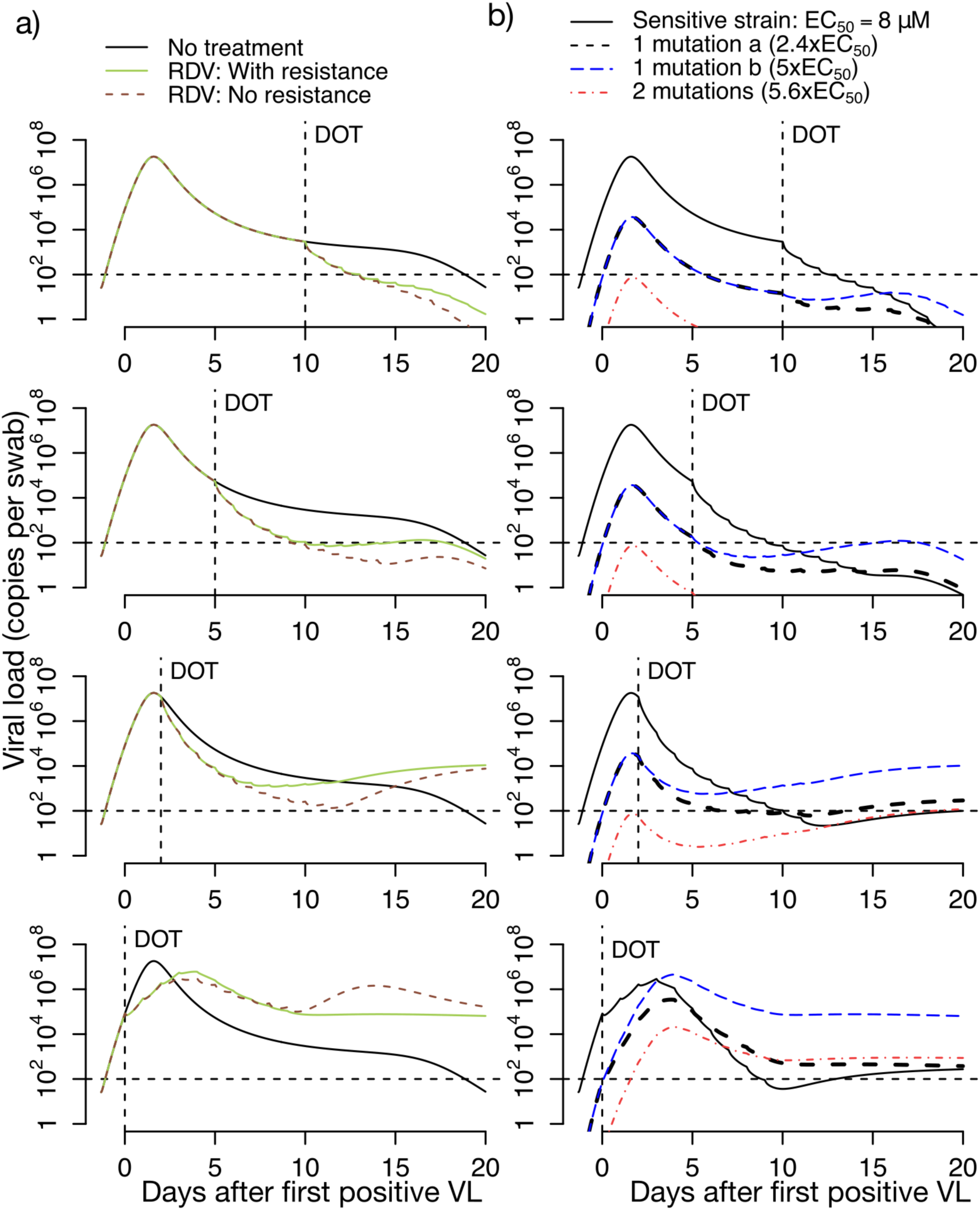
Projections of remdesivir drug resistance during therapy. Simulations are with moderate potency (EC50=8.0 uM) and the assumption that mutants confer partial drug resistance. Treatment initiation is at timepoints generally consistent with hospitalization (day 10 after first positive sample), first symptoms (day 5 after first positive sample), pre-symptomatic post-peak phase (day 2 after first positive sample) or pre-symptomatic pre-peak phase (day 0). **A**. Projections of no treatment, treatment with no assumed drug resistance, and treatment with assumed drug resistance. **B**. Projections of assumed drug resistance with trajectories of sensitive strains, single mutants and double mutants.

If we assume that a single point mutation in SARS CoV-2 could theoretically induce complete resistance, then treatment failure may occur regardless of dose timing. The resistant strain is predicted to predominate raising the possibility of transmitted drug resistance **(Fig. S2)**.

### Hydroxychloroquine treatment predictions

Using a multi-compartment PK/PD model for hydroxychloroquine **(Fig. S3a)**, we first recapitulated drug levels of the drug over time following a single dose **(Fig. S3b)**, simulated twice daily doses over 10 days **(Fig. S3c)**, generated theoretical dose response curves **(Fig. S3d)**, and then projected drug efficacy over time **(Fig. S3e)**. As with remdesivir, the predicted efficacy of therapy on shedding depended on timing of treatment and the intracellular *in vivo* EC50 **(Fig. S4a-e)**. Lowering the area under the curve again required extremely early initiation of potent treatment **(Fig. S4f)** while decreasing the number of effector cells required effective dosing within the first 5 days of infection **(Fig. S4g)**.

### Broadly neutralizing antibody predictions

We next simulated a single infusion of broadly neutralizing antibodies with prolonged half-life. Broadly neutralizing antibodies are designed to stop infection of new cells rather than eliminate viral replication. We used a dual compartment PK model for **(Fig. S5a)**, estimated antibody levels over time following a single dose **(Fig. S5b)**, generated theoretical dose response curves **(Fig. S5c)**, and then projected antibody efficacy over time **(Fig. S5d)**. The predicted efficacy of therapy on shedding again was dependent on timing of treatment and the *in vivo* EC50 **(Fig. S6a-d)**. Once again, to lower duration of shedding and the number of effector cells depended on therapeutic potency, while effective lowering of viral area under the curve required extremely early initiation of potent treatment **(Fig. S6e-g)**.

### Immunotherapy predictions

We generalized the potential effects of a cellular immunotherapy which would presumably decrease the lifespan of infected cells. We projected that such an intervention would need to increase this rate 10-fold to match the efficacy of a potent small molecular agent. **(Fig. S7a-c)**. Immunotherapies were projected to have no efficacy if started prior to peak shedding **(Fig. S7d)**.

## Discussion

SARS CoV-2 infection is characterized by a stereotypical viral kinetic pattern with a high peak viral load during the first several days of infection, a subsequent short rapid decay period followed by a slower clearance phase of variable duration, and a final, rapid elimination phase. Our mathematical model reproduces this data and explains that the transition from first to second phase viral decay is governed by a density dependent term in which a massive die off of infected cells occurs during peak viremia. Viral clearance rate slows considerably once the viral load decreases below a threshold of ∼10^5^ RNA copies. A late slowly expanding cytolytic T cell response is predicted to induce eradication of infected cells in the upper airway 2-20 days later. The timing of this final response appears to be variable among infected people. Moreover, it is unknown if cessation of viral replication in the nasopharynx guarantees the same result in the lung or other anatomic compartments (*8*).

We built therapy simulations on top of these natural kinetics to glean knowledge about the possible impact of antiviral therapies. While accurate prediction is impossible in the absence of clinical trial data, we are able to make several observations which highlight the importance of off treatment viral dynamics in understanding treatment effects. Our results can guide future study design and assist in interpretation of forthcoming trial data in multiple ways.

First, it is critical to know whether *in vitro* potency assessments of remdesivir, hydroxycholorquine and bNAbs can be projected onto human infection. We previously demonstrated that *in vivo* IC50 values for antiviral agents can exceed estimates derived from cell cultures experiments by a multiple of 5-10 for small molecule antiviral drugs (*19*). A similar observation has been hypothesized for HIV targeting monoclonal neutralizing antibodies (*23*). It is unclear if this discrepancy occurs due to low blood levels in tissue, different cell metabolism of drug in tissue or higher protein binding *in vivo*. Whatever the case, if *in vitro* potency measurements of hydroxycholoroquine and remdesivir overestimate *in vivo* activity, or if higher intracellular levels are required than in plasma, then these drugs may be less effective in clinical trials. Higher dosing may be a possible solution to circumvent this issue.

Second, effective dosing after symptom development predicts rapid subsequent elimination of infected. Most current clinical trials are focused primarily on hospitalized patients whereas our results and those from other COVID models suggest that treatment in the days immediately following symptom onset will decrease the duration of detectable viral shedding (*24, 25*). Our model also predicts that early treatment will limit the extent of the cytolytic immune response required to clear infection. If either of these outcomes are correlates of progression to severe disease and transmission risk, then as with HIV (*26*), influenza (*27*), and Ebola (*28*), early test and treat is a vital, currently overlooked strategy.

Third, effective dosing soon after onset of symptoms is predicted to have an insignificant effect on viral load AUC. This phenomenon occurs because the amount of virus produced per hour at the early peak is far higher than the amount produced per day during the lower viral load second decay phase. This finding provides a cautionary message: if subsequent disease severity and development is imprinted during the high viral load, pre-symptomatic phase of infection, then early antiviral therapy could theoretically eliminate further shedding without altering clinical outcomes. Moreover, if extremely high viral load periods are responsible for most transmissions as suggested by the 5-day generation time of infection (*29*), then early elimination of shedding may not lower transmissibility either. Overall this result highlights the urgent need for studies which identify early viral and immune correlates of severe disease and transmissibility. Indeed, preliminary studies suggest that presenting viral load may impact disease severity (*30*).

Fourth, if therapy is initiated during the pre-symptomatic stage of infection as may occur during post exposure prophylaxis (*31*), then its effectiveness is predicated on high potency.

Subtherapeutic small molecular agents may in fact prolong infection. Based on this result, we suggest sampling at late timepoints during post exposure prophylaxis trials.

Fifth, our approach suggests that while development of low-level drug resistance will occur commonly during COVID-19 treatment with remdesivir, it is unlikely to predispose to treatment failure provided the drug is potent against the predominant susceptible strain. Resistant variants will likely be present at low levels relative to susceptible strains making transmission of resistance far less likely. Important exceptions may occur in immunocompromised hosts who might shed respiratory viruses for longer and at higher levels, thereby increasing the chance of *de novo* resistance (*32*), or in the context of only moderately potent antiviral therapy.

Finally, our model projects a high likelihood of success for neutralizing antibodies and cellular immunotherapies provided that they achieve adequate potency and are dosed early.

In summary, our model provides a broad platform for assessment of all major types of therapies. Our results demonstrate the need to differentiate whether duration of viral shedding or viral area under the curve is the more relevant surrogate of severity. If viral area under the curve is most predictive of poor outcomes, then all forms of antiviral therapy outside of potent post exposure prophylaxis are unlikely to provide clinical benefit. However, if shedding duration is the best surrogate, then an early test and treat approach is highly promising for limiting the likelihood of severe disease.

## Materials and Methods

### Study design

We employed ordinary differential equation models to analyze the in-host SARS CoV-2 dynamics in infected individuals and the potential *in vivo* effect of different treatment strategies. First, we fit models to the viral load data from different sources using a nonlinear-mixed effects approach. Second, we used pharmacokinetics models to fits observed plasma concentration of remdesivir (RDV) and its active nucleoside triphosphate form in PBMCs, and blood concentration of hydroxychloroquine (HCQ). Third, we simulated dose response curves for antiviral effect of RDV and HCQ using different possible half maximal effective concentration (EC50) based on *in-vitro* estimations against SARS-CoV-2. Fourth, we simulated therapy at different times during infection to analyze the potential reduction of SARS CoV-2 shedding. Finally, we repeated projections of therapy including the emergence of resistance to therapy.

### SARS CoV-2 viral load data

We analyzed viral load data from patients infected with SARS CoV-2 that were monitored and received supported therapy in hospitals in Singapore (n=11), Germany (n=9), South Korea (n=1), and France (n=4). Patients who had less than 4 data points or had oscillatory viral dynamics were excluded.

The first data set was obtained from SARS Cov-2-infected patients followed at 4 hospital in Singapore from January 23^rd^ to February 23^rd^, 2020 (*7*). All patients diagnosed stated to have travelled from Wuhan, China in the last two weeks of enrollment. Viral load observations were obtained from different specimens (blood, stool, and urine samples), but we analyzed those coming from nasopharyngeal swabs. Cycle threshold were obtained with reverse transcriptase polymerase chain reaction (RT-PCR) at multiple times during the first 2 weeks after enrollment.

The second data set was obtained from infected patients enrolled and treated in a single hospital in Munich, Germany from January 23^rd^ to January 27^th^, 2020 (*8*). For all patients, the infection was reported to happen after contact with an index case. Viral load observations were obtained daily from sputum, pharyngeal swabs and stool using RT-PCR. Here, we analyzed viral RNA concentrations from sputum.

The third data set comes from the first SARS Cov-2 infected case in Korea, a 35-year-old Chinese citizen coming from Wuhan, China (*9*). Nasopharyngeal swabs viral loads were obtained daily from day 2 of symptoms onset using RT-PCR.

The fourth data set came from four patients admitted in hospitals in Paris or Bordeaux, France with viral loads obtained from nasopharyngeal swabs using RT-PCR (*10*). We analyzed viral load data digitized from the published study.

When viral load observations were only published in cycle threshold (Ct) values we converted them to copies per swab using the relation values in (*33*). We assumed a lower limit of detection of 100 copies per sample.

### Pharmacokinetic data

PK data of HCQ specifically to 200 mg oral dose was gathered from (*34*). PK data of RDV was gathered from (*17*), where non-Human Primates (NHPs) intravenously received RDV at 10mg/kg dose at day 0 and the plasma concentration of RDV and its active nucleoside triphosphate form in PBMCs were recorded over 24 hours. We digitize this data and employ it to fit PK model of RDV.

### Mathematical modeling of SARS-COV-2 dynamics

To understand the observed SARS-COV-2 shedding dynamics we developed a viral infection model modifying previous models of virus dynamics (*12, 35-37*). In this model, susceptible cells (*S*) are infected at rate *βVS* by SARS-COV-2 (*V*). SARS-COV-2-infected cells (*I*) are cleared in two ways: (1) by an innate response with density dependent rate *δI*^*k*^(*13, 14*); and (2) an acquired response with rate 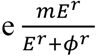 mediated by SARS-COV-2-specific effector cells (*E*). The exponent *k* describes by how much the first death rate depends on the infected cell density. The Hill coefficient *r* parameterizes the nonlinearity of the second response and allows for rapid saturation of the killing. Parameter *ϕ*defines the effector cell level by which killing of infected cells by *E* is half maximal. SARS-COV-2 is produced at a rate *π*and cleared with rate *γt* In the model, SARS-COV-2-specific effector cells rise after *n* stages from precursors cells (*M*_*i*=1…*n*_). The first precursor cell compartment (*M*_1_) proliferates in the presence of infection with rate *ωIM*_1_and differentiates into the effector cell at a per capita rate *q* during each intermediate stage. Finally, effector cells die at rate *δ*_*E*_. The best instance of the model is expressed as a schematic (**Fig. 1a**) and here as a system of ordinary differential equations:

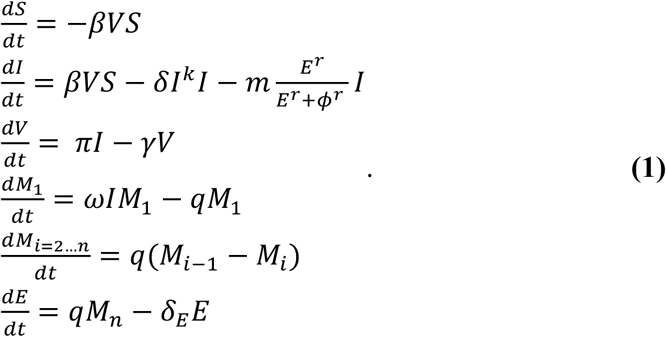

### Fitting viral load data and model selection

We fit different instances of our model in **equation 1** to the SARS-COV-2 shedding data using a nonlinear mixed-effects modeling approach (*38-40*) (See **Table S1**). Briefly, we obtained a maximum likelihood estimation of the population median (fixed effects) and standard deviation (random effects) for each model parameter using the Stochastic Approximation Expectation Maximization (SAEM) algorithm embedded in the Monolix 2019R2 software (www.lixoft.eu). For a subset of parameters, random effects were specified, and the standard deviation values were estimated. Measurement error variance was also estimated assuming an additive error model for the logged *V*. We simultaneously fit each model to the viral load data of 25 patients form the four data sets. The parameters associated with the effector cell compartment were only estimated for those study participants who cleared infection during the observed data.

For each model fit we assumed *t* =0 as the time of first positive viral load for each person. However, we defined the initial value as the time of infection, i.e. when *I*(*t*_*init*_)=1cell. Since infection starts before the first detected viral load, we have that *t*_*init*_ <0. We fixed other initial values as *S*(*t*_*init*_)= 10^7^cells, 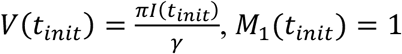 cell and *M*_*i*=2…*n*_(*t*_*init*_)= *E*(*t*_*init*_)=0. We fixed parameter *γ* =15day^-1^ (*41*), *δ*_*E*_=1day^-1^ (*35*), and *ϕ*= 100cells. We assumed this value of *ϕ*because of the low percentage (∼2%) of activated T cells that start growing at the moment of viral load drop (*42*). We estimated the remaining parameters including the time of infection *t*_*init*_.

To determine the most parsimonious model among the instances for the available SARS Cov-2 shedding data, we computed the log-likelihood (log *L*) and the Akaike Information Criteria (AIC=-2log *L*+2*m*, where *m* is the number of parameters estimated). We assumed a model has similar support from the data if the difference between its AIC and the best model (lowest) AIC is less than two (*43*).

### Pharmacokinetic modeling of remdesivir

To reproduce the PK data of RDV, we employed a simple two compartment model where the first compartment represents the amount of RDV in plasma (*C*_*P*_, volume *V*_*P*_) and the second compartment denotes the amount of its active nucleoside triphosphate form in PBMCs (*C*_*a*_, volume *V*_*a*_). Here, we assume that RDV gets metabolized to its active nucleoside triphosphate form at rate *k*_*pa*_ whereas RDV and its active nucleoside triphosphate form are eliminated from their respective compartments at rates *k*_*c*;_ and *k*_*a*_, respectively. The model is given by,

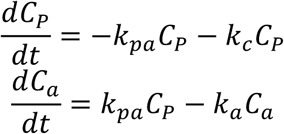

As the drug concentration was recorded in *μ*M, we convert the amount of the drug given in mg to *μ*M by dividing *C*_*P*_ and *C*_*a*_ by conversion factors *V*_1_and *V*_2_, respectively. Here, *V*_1_and *V*_2_ are a combination of the volume of the distribution of two compartments and the molar mass of two forms of RDV.

We fit this model to the PK data from NHP (*17*), using a non-linear least squares approach using an initial dose of 10mg/kg. We adapted the dose in humans (200mg and 100mg) assuming a normal weight of 70kg.

### Pharmacokinetic modeling of hydroxychloroquine

To recapitulate the pharmacokinetics of HCQ, we developed a model on pharmacokinetics models of chloroquine (CQ), as HCQ is a derivate of CQ and has the same active metabolites (*44*). In this four-compartment model, the drug gets absorbed from the gut compartment (*A*_*G*_) at rate *k*_*a*_ and enters the central compartment (*A*_*P*_, with volume *V*_1_), where it is eliminated with at a rate *k*_*c*_. The rate constants *k*_12_ and *k*_13_represent the movement of the drug from the central compartment to the first peripheral compartment (*A*_1_, volume *V*_1_) and the first peripheral compartment (*A*_3_, volume *V*_3_), respectively. Similarly, the movement of the drug from compartments *A*_1_ and *A*_3_to the central compartment is modelled using *k*_21_and *k*_31_, respectively. The modeling equations representing the amount of drug in each compartment is given by,

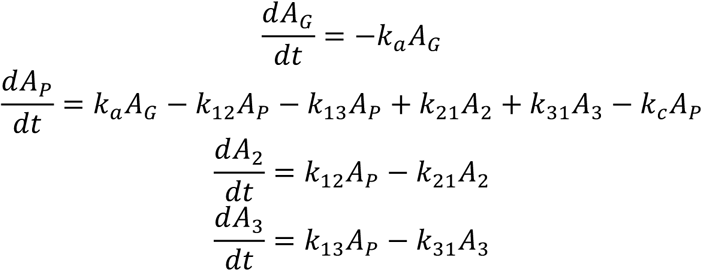

We fit this model to the PK data using a non-linear least squares approach.

### Pharmacokinetic modeling of broadly neutralizing antibodies

The pharmacokinetics of a intravenously injected broadly neutralizing antibody (bNAB) was simulated using a simple bi-phasic exponential model,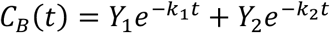 (*45*). In this model *k*_1_ad *k*_2_ represent the distribution and clearance rates of the bNAB whereas *Y*_1_and *Y*_2_ describe the coefficients associated with them. We fixed parameters *Y*_1_=2200*μg*/mL, *Y*_2_ =150*μg*/mL and *k*_1_=1.1day^-1^ following estimates for the bNAb VRC01 (*23*). We also fixed 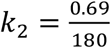 day^-1^ to reflect the long half-life of 3 months.

### Pharmacodynamic modeling

We model antiviral efficacy (*∈*) of each treatment approach as a function of the drug concentration *C*(*t*)as 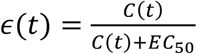 being *EC*_50_ the half maximal effective concentration. Remdesivir and hydroxychloroquine are assumed to inhibit viral production rate (*π*) whereas bNABs are hypothesized to inhibit the viral infectivity (*β*), both by a factor of 1−*∈*(*t*).

To calculate the efficacy of remdesivir, we assume that the active form (*C*_*a*_)has the antiviral effect, i.e.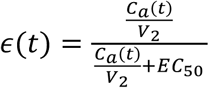. Since concentration of HCQ in lung and other tissues reaches levels of 200-700 times higher than in plasma (*46*), we took a conservative approach and determined an intracellular concentration of HCQ as 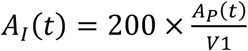.Thus, the antiviral effect of intracellular HCQ was calculated as 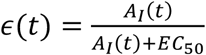. We used values of EC50 from those estimates *in vitro* against SARS CoV-2 (0.72 μM for Hydroxychloroquine (*47*), and 0.77 μM for Remdesivir (*16*)) to hypothetical *in* vivo values up to 100 times the *in vitro* value.

We also explored the antiviral effect of a hypothetical bNAb using the form 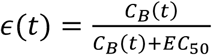 and varied values of EC50 between 1 and 1000 μM. Finally, we modeled the potential antiviral effect of immunotherapies by enhancing the early death rate of infected cells (*δ*) by a factor of 50%, 100% and 200%.

### Modeling the emergence of resistance to remdesivir

We adapted the model in **equation 1** to allow for the emergence of resistance to RDV. We base the modifications on the fact that two separate mutations may induce partial resistance to RDV in SARS CoV-1 which in turn leads to a less fit virus (*22*). In the case of SARS Cov-1, two single mutations can induce to a less sensitive virus with 2.4- and 5.0-fold increase in the RDV EC50. When the combined mutations emerged, it mediated a 5.6-fold increase in the RDV EC50. We included these findings in the model by assuming that infected cells (*I*_*s*_) that produce sensitive virus (*V*_*s*_) can transition into infected cells (*I*_*r*1_and *I*_*r*2_) that produce less sensitive virus (*V*_*r*1_and *V*_*r*2_), due to one mutation during the viral replication cycle. These two viral populations have an increased EC50 (2.4- and 5-fold higher). Similarly, we assumed that *I*_*r*1_and *I*_*r*2_ can transition into infected cells that produce a more resistant strain *V*_*r*12_ (with 5.6-fold higher EC50) after another mutation. We also allowed for reversal mutation events. We assumed a mutation probability of *μ* = 10^*−5*^per infection event. Under these assumptions total viral load is defined as *V* = *V*_*s*_ *+ V*_*r*1_*+ V*_*r*2_ *+ V*_*r*12_ and total number of infected cells as *I* = *I*_*s*_ *+ I*_*r*1_*+ I*_*r*2_ *+ I*_*r*12_. With these modifications the model becomes,

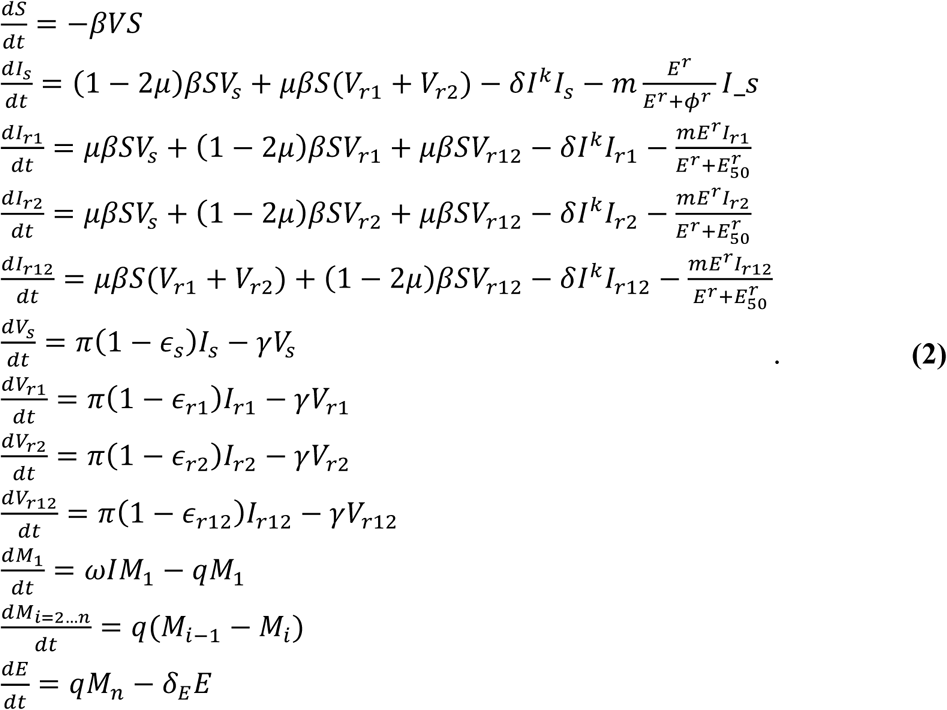

Here,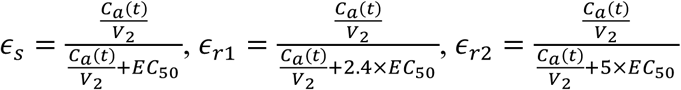 and 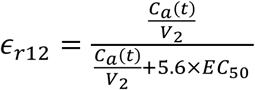 are the antiviral effect of Remdesivir in blocking virus production for each viral population.

## Data Availability

All data and code will be made available upon request.

## Author contributions

J.T.S. conceived the study. A.G. and E.F.C. assembled data, wrote all code, performed all calculations and derivations, ran the models, and analyzed output data. J.T.S. wrote the manuscript with contributions from all other authors.

## Data and materials availability

Original data and code will be shared upon request.

---

**Fig S1.**
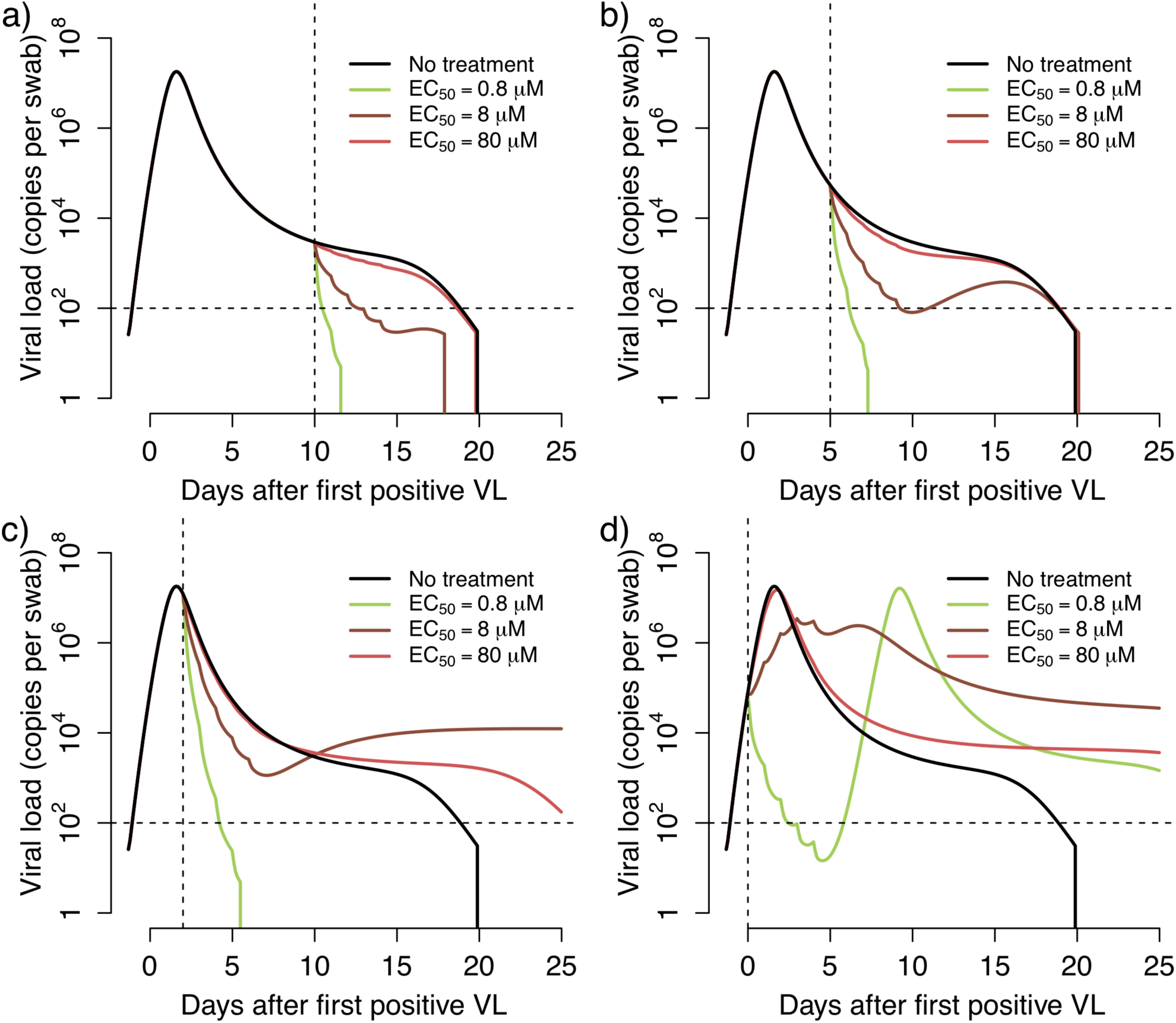
Treatment projections of a 5-day remdesivir course assuming different potency and timing of treatment. Each set of simulations is performed under assumptions of high, medium and low potency (EC50=0.8, 8 and 80 uM respectively). Treatment initiation at timepoints generally consistent with **a**. hospitalization (day 10 after first positive sample), **b**. first symptoms (day 5 after first positive sample), **c**. pre-symptomatic post-peak phase (day 2 after first positive sample) and **d**. pre-symptomatic pre-peak phase (day 0). Overall. Early potent treatment limits duration of infection.

**Fig S2.**
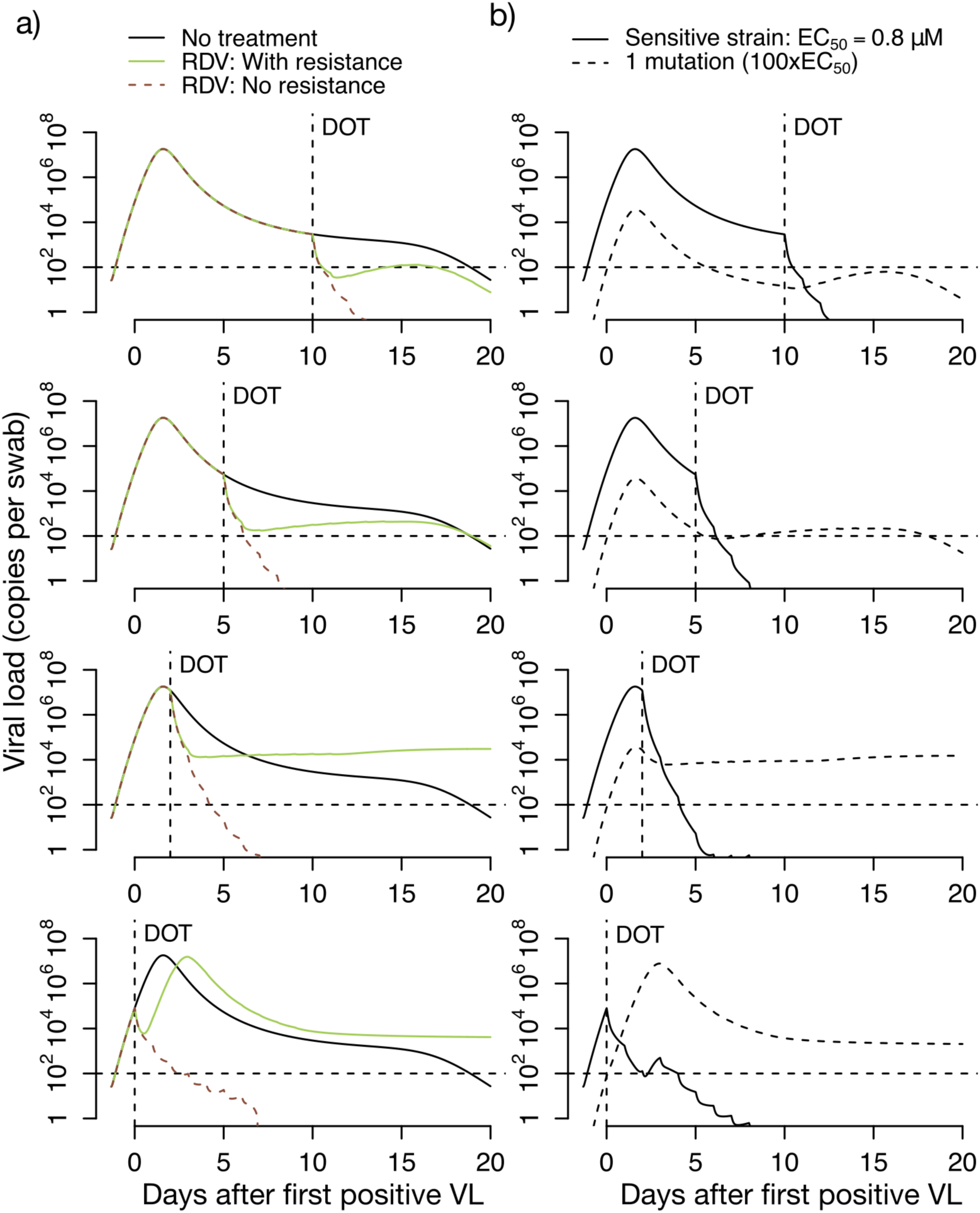
Projections of remdesivir drug resistance during therapy. Simulations are with high potency (EC50=0.8 uM) and the assumption that mutants confer complete drug resistance. Treatment initiation is at timepoints generally consistent with hospitalization (day 10 after first positive sample), first symptoms (day 5 after first positive sample), pre-symptomatic post-peak phase (day 2 after first positive sample) or pre-symptomatic pre-peak phase (day 0). **A**. Projections of no treatment, treatment with no assumed drug resistance, and treatment with assumed drug resistance. **B**. Projections of assumed drug resistance with trajectories of sensitive strains and single mutants.

**Fig S3.**
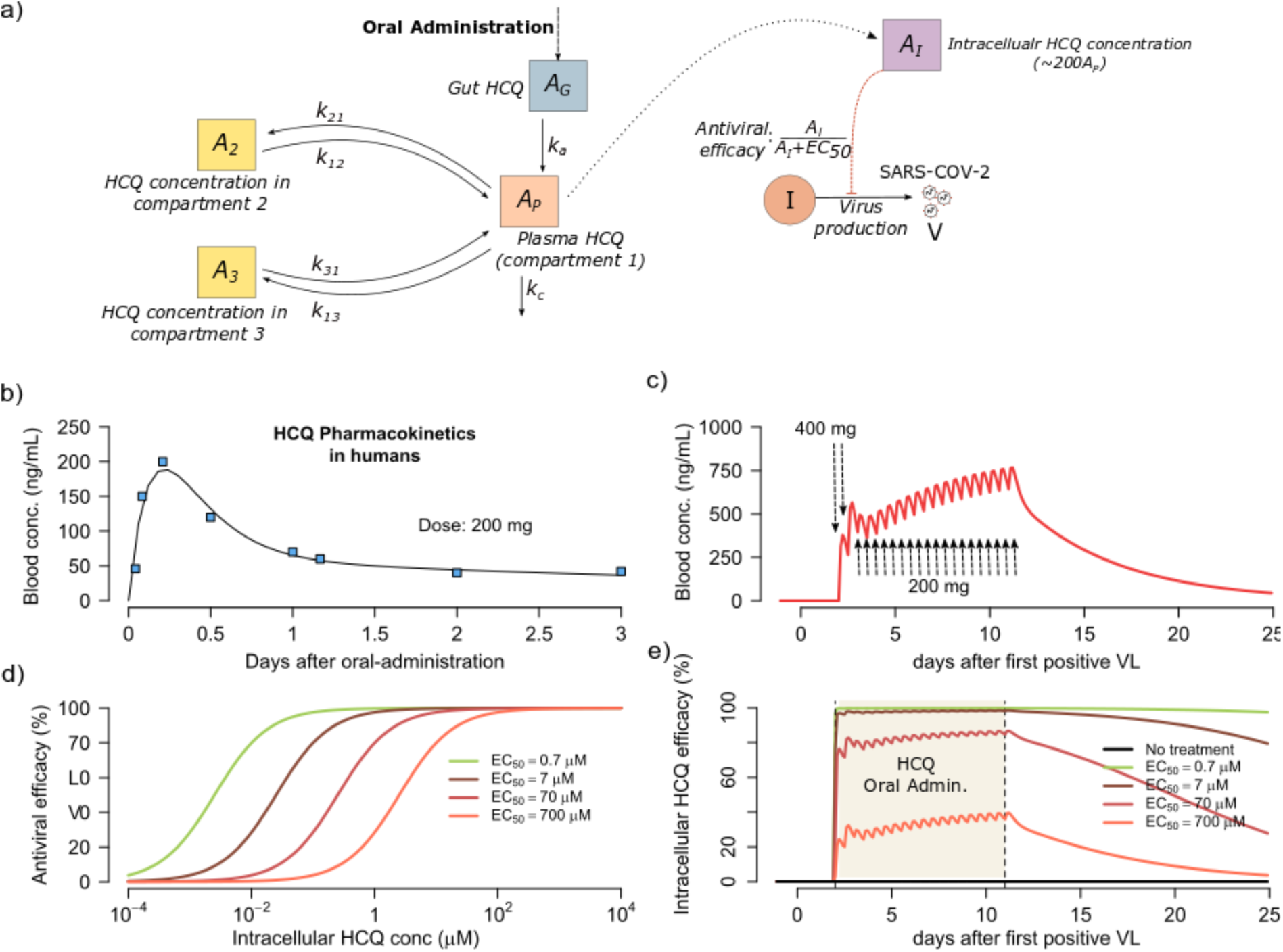
Projected pharmacokinetics and pharmacodynamics of hydroxychloroquine therapy. **a**. Complete model of hydroxychloroquine (HCQ) including gut, plasma and dual compartment levels of parent drug, intracellular levels of the active component and antiviral efficacy of drug according to concentration. **b**. Projections of plasma HCQ levels. Datapoints from human PK experiments are dots while lines are model projections. **c**. Simulated concentrations of the parent compound with a loading dose of twice daily 400 mg followed by 18 twice daily doses of 200 mg. **d**. Pharmacodynamic projections of antiviral efficacy according to drug concentration assuming different values for the *in vivo* EC50 of the drug. **e**. Combination simulations of pharmacokinetic and pharmacodynamic models demonstrating prolonged antiviral activity after dosing is stopped.

**Fig S4.**
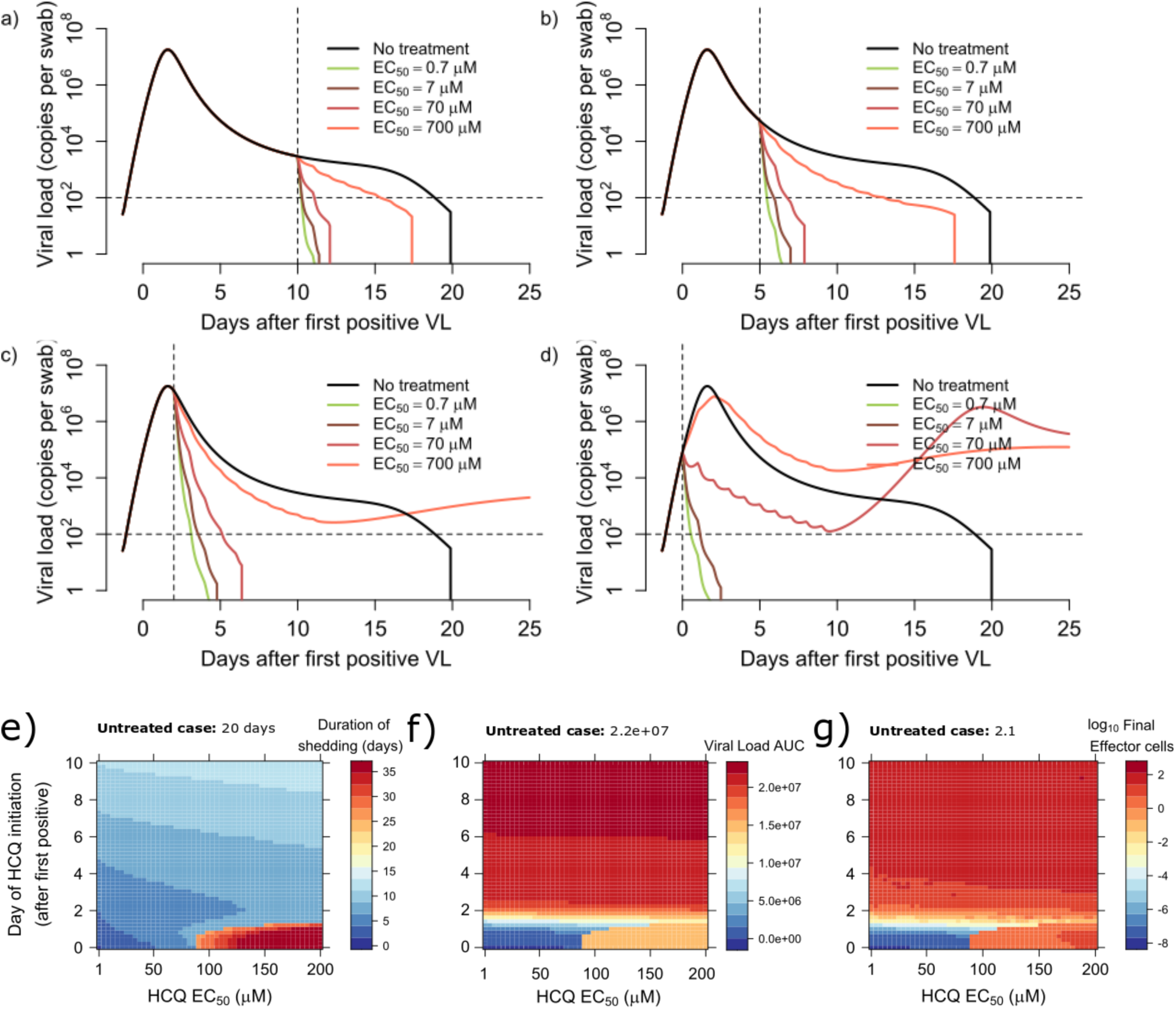
Treatment projections of hydroxychloroquine assuming different potency and timing of treatment. Each set of simulations is performed under assumptions of high, medium and low potency. Treatment initiation is at timepoints generally consistent with **a**. hospitalization (day 10 after first positive sample), **b**. first symptoms (day 5 after first positive sample), **c**. pre-symptomatic post-peak phase (day 2 after first positive sample) and **d**. pre-symptomatic pre-peak phase (day 0). Overall, early potent treatment limits duration of infection. Heatmaps comparing variance in drug potency measured by *in vivo* EC50 (x-axis) and timing of treatment initiation (y-axis) for **e**. Shedding duration, **f**. viral load area under the curve (AUC) and **g**. extent of T cell response required for viral elimination.

**Fig S5.**
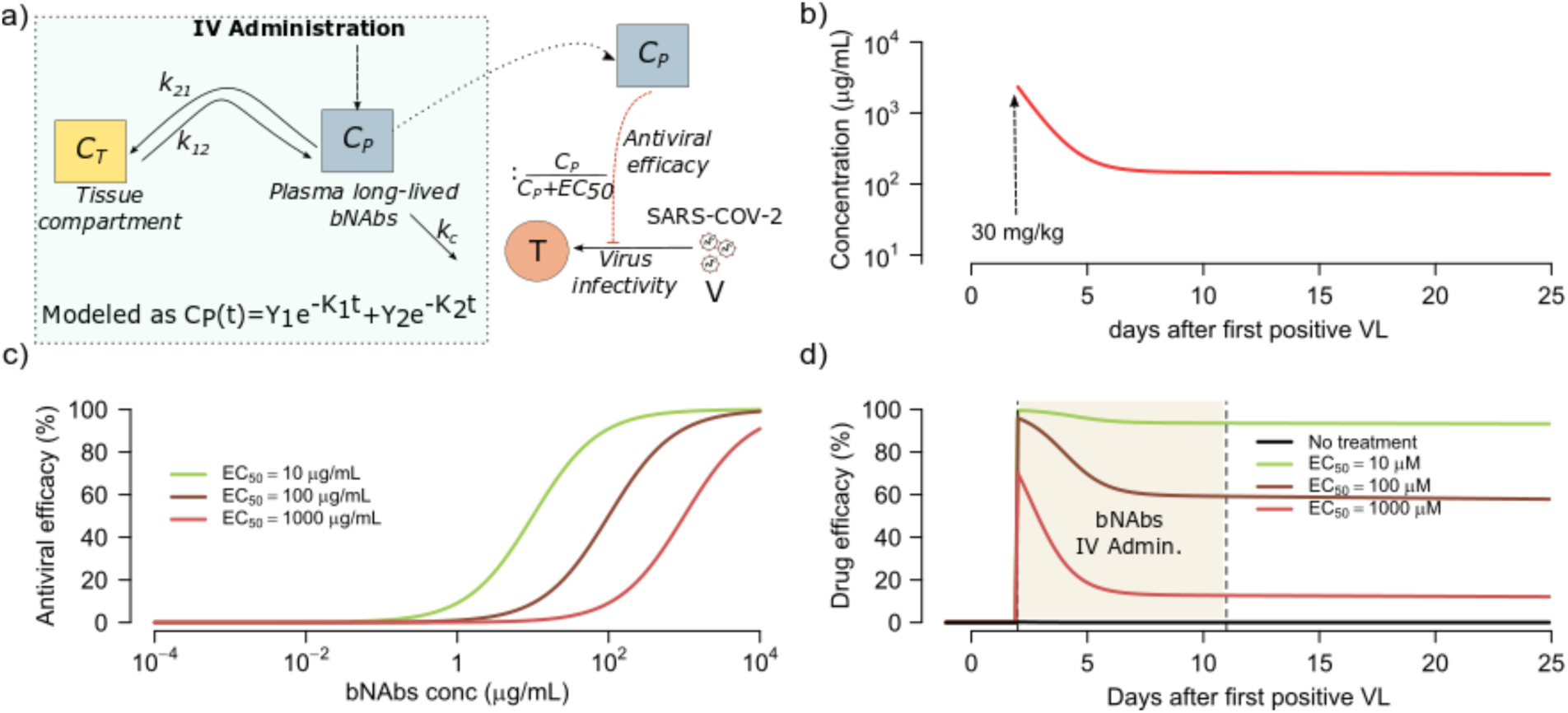
Projected pharmacokinetics and pharmacodynamics of neutralizing antibody therapy. **a**. Complete bi-compartment model of broadly neutralizing antibody therapy with lowering of viral infectivity according to antibody concentration. **b**. Projections of plasma bNAb levels from simulations of VRC01 treatment **c**. Pharmacodynamic projections of antiviral efficacy according to antibody concentration assuming different values for the *in vivo* EC50. **d**. Combination simulations of pharmacokinetic and pharmacodynamic models demonstrating antiviral activity as a function of time

**Fig S6.**
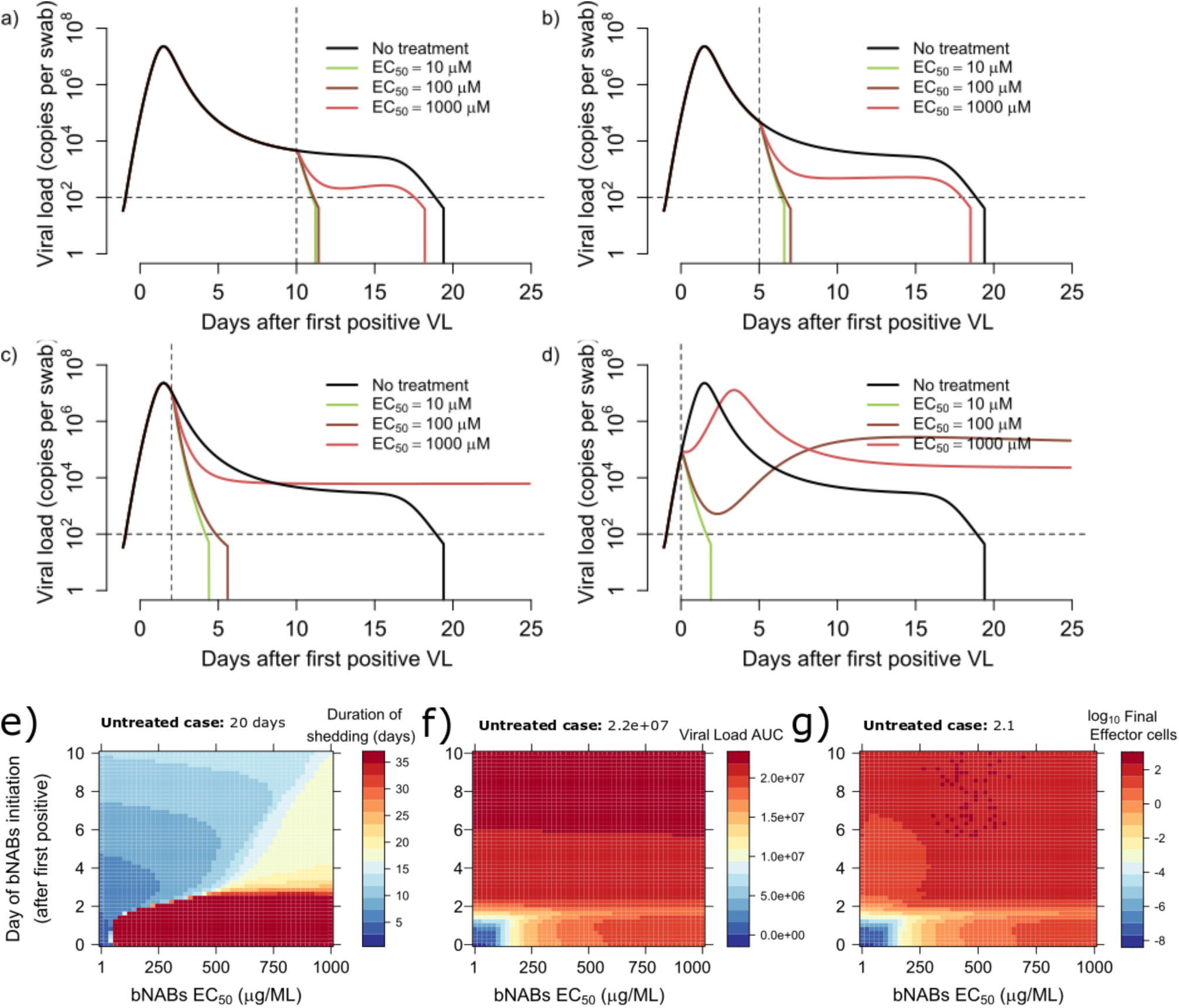
Treatment projections of broadly neutralizing antibody (bNab) assuming different potency and timing of treatment. Each set of simulations is performed under assumptions of high, medium and low potency. Treatment initiation is at timepoints generally consistent with **a**. hospitalization (day 10 after first positive sample), **b**. first symptoms (day 5 after first positive sample), **c**. pre-symptomatic post-peak phase (day 2 after first positive sample) and **d**. pre-symptomatic pre-peak phase (day 0). Overall, early potent treatment limits duration of infection. Heatmaps comparing variance in bNAb potency measured by *in vivo* EC50 (x-axis) and timing of treatment initiation (y-axis) for **e**. Shedding duration, **f**. viral load area under the curve (AUC) and **g**. extent of T cell response required for viral elimination.

**Fig S7.**
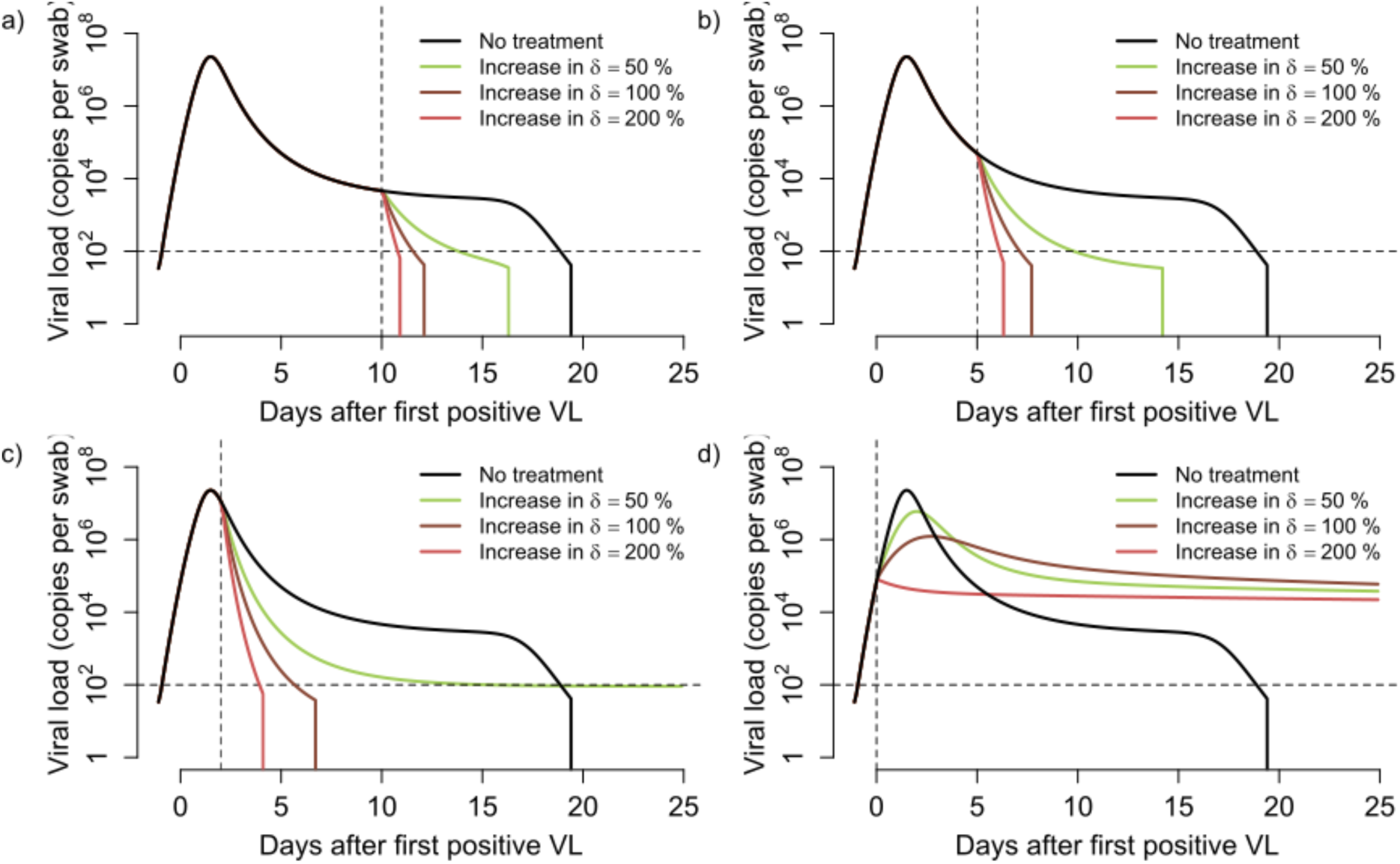
Treatment projections of cytolytic immunotherapy assuming different potency and timing of treatment. Each set of simulations is performed under assumptions of high, medium and low potency based on multiplicative effect on infected cell death rate. Treatment initiation is at timepoints generally consistent with **a**. hospitalization (day 10 after first positive sample), **b**. first symptoms (day 5 after first positive sample), **c**. pre-symptomatic post-peak phase (day 2 after first positive sample) and **d**. pre-symptomatic pre-peak phase (day 0). Overall, early potent treatment limits duration of infection.

**Table S1:** Akaike information criterion (AIC) for multiple instances of our model with different number of the compartments of *M*(*n*) and the hill-coefficient associated the effector cell response (*r*). Lower the AIC, better is the support for the combination of parameters for our model. We found that *n*=2and *r*= 10 is best supported by the data (bold red). For comprehensive, we also tried a model with *n*=0 (i.e., no effector cell response), and a model with *n*=0 and *k*=0. In both cases, we found AIC∼945, supporting the choice of our model.

| | $r = 0.1$ | $r = 1$ | $r = 5$ | $r = 10$ | $r = 15$ |
| --- | --- | --- | --- | --- | --- |
| $n = 1$ | 922.8 | 935.4 | 929.7 | 926.5 | 932.2 |
| $n = 2$ | 929.2 | 924.0 | 941.6 | **919.9** | 925.9 |
| $n = 3$ | 922.8 | 935.4 | 929.7 | 926.5 | 932.2 |
| $n = 4$ | 951.5 | 928.6 | 925.8 | 925.5 | 924.3 |
| $n = 5$ | 950.4 | 930.8 | 929.1 | 923.3 | 922.5 |
| $n = 6$ | NA | 928.6 | 926.0 | 924.9 | 923.2 |
| $n = 7$ | NA | 928.3 | 926.4 | 927.8 | 924.8 |

**Table S2.** Individual parameter estimates for the best model fits to the viral load data (lowest AIC in Table S1).

| **ID** | **$t_{init}$**<br>(days<br>before<br>1<sup>st</sup> +) | **$\log_{10}\beta$**<br>(virions<sup>-1</sup><br>day<sup>-1</sup>) | **$\delta$**<br>(day<sup>-1</sup><br>cells<sup>-k</sup>) | **$k$**<br>(-) | **$\log_{10}\pi$**<br>(log<sub>10</sub> day<sup>-1</sup>) | **$m$**<br>(day<sup>-1</sup><br>cells<sup>-1</sup>) | **$\log_{10}\omega$**<br>(day<sup>-1</sup><br>cells<sup>-1</sup>) | **$q$**<br>(day<sup>-1</sup>) |
| --- | --- | --- | --- | --- | --- | --- | --- | --- |
| **S2** | -6.68 | -7.23 | 3.14 | 0.08 | 2.59 | 2.91 | -4.56 | 2.4E-05 |
| **S3** | -3.73 | -7.22 | 3.12 | 0.08 | 2.60 | 3.21 | -4.55 | 2.4E-05 |
| **S4** | -6.56 | -7.23 | 3.14 | 0.08 | 2.59 | 3.51 | -4.56 | 2.4E-05 |
| **S5** | -2.00 | -7.23 | 3.15 | 0.08 | 2.59 | 3.31 | -4.56 | 2.4E-05 |
| **S6** | -2.79 | -7.23 | 3.12 | 0.08 | 2.60 | 3.21 | -4.55 | 2.4E-05 |
| **S10** | -5.66 | -7.23 | 3.13 | 0.08 | 2.59 | 3.54 | -4.55 | 2.4E-05 |
| **S11** | -4.80 | -7.23 | 3.13 | 0.08 | 2.60 | 3.51 | -4.56 | 2.4E-05 |
| **S12** | -6.29 | -7.23 | 3.14 | 0.08 | 2.59 | 3.26 | -4.56 | 2.4E-05 |
| **S14** | -1.23 | -7.23 | 3.13 | 0.08 | 2.59 | 3.57 | -4.55 | 2.4E-05 |
| **S17** | -7.06 | -7.23 | 3.14 | 0.08 | 2.59 | 3.39 | -4.56 | 2.4E-05 |
| **S18** | -1.32 | -7.23 | 3.15 | 0.08 | 2.59 | 3.20 | -4.56 | 2.4E-05 |
| **G1** | -1.27 | -7.22 | 3.12 | 0.08 | 2.60 | 3.21 | -4.55 | 2.4E-05 |
| **G2** | -0.92 | -7.22 | 3.12 | 0.08 | 2.60 | 3.21 | -4.55 | 2.4E-05 |
| **G3** | -4.57 | -7.22 | 3.12 | 0.08 | 2.60 | 3.21 | -4.55 | 2.4E-05 |
| **G4** | -3.97 | -7.23 | 3.12 | 0.08 | 2.60 | 3.13 | -4.55 | 2.4E-05 |
| **G5** | 1.32 | -7.22 | 3.12 | 0.08 | 2.60 | 3.21 | -4.55 | 2.4E-05 |
| **G6** | -2.76 | -7.23 | 3.13 | 0.08 | 2.60 | 3.21 | -4.55 | 2.4E-05 |
| **G7** | -2.00 | -7.23 | 3.13 | 0.08 | 2.59 | 3.21 | -4.55 | 2.4E-05 |
| **G8** | -6.12 | -7.23 | 3.12 | 0.08 | 2.60 | 3.21 | -4.55 | 2.4E-05 |
| **G9** | -7.58 | -7.22 | 3.11 | 0.08 | 2.60 | 3.72 | -4.55 | 2.4E-05 |
| **K1** | -4.17 | -7.23 | 3.14 | 0.08 | 2.59 | 3.30 | -4.56 | 2.4E-05 |
| **E1** | -4.02 | -7.22 | 3.10 | 0.08 | 2.60 | 4.06 | -4.55 | 2.4E-05 |
| **E3** | -4.14 | -7.23 | 3.14 | 0.08 | 2.59 | 3.36 | -4.56 | 2.4E-05 |
| **E4** | -3.74 | -7.23 | 3.11 | 0.08 | 2.60 | 3.45 | -4.55 | 2.4E-05 |
| **E5** | -2.31 | -7.23 | 3.11 | 0.08 | 2.60 | 3.54 | -4.55 | 2.4E-05 |

**Table S3.** Individual parameter estimates for pharmacokinetics model of remdesivir.

| **Parameter** | **Value** |
| --- | --- |
| $k_{pa}$ | 0.86 day<sup>-1</sup> |
| $k_c$ | 1.2 day<sup>-1</sup> |
| $k_a$ | 0.05 day<sup>-1</sup> |
| $V_1$ | 2.84 |
| $V_2$ | 0.12 |

**Table S4.** Parameter estimates for pharmacokinetics model of hydroxychloroquine.

| Parameter | Value |
| --- | --- |
| $k_a$ | 6.1 day<sup>-1</sup> |
| $k_c$ | 229 day<sup>-1</sup> |
| $k_{12}$ | 2*10<sup>6</sup> day<sup>-1</sup> |
| $k_{21}$ | 1.5*10<sup>4</sup> day<sup>-1</sup> |
| $k_{13}$ | 6.5*10<sup>2</sup> day<sup>-1</sup> |
| $k_{31}$ | 0.85 day<sup>-1</sup> |
| $V_1$ | 0.002 mL |
| $V_2$ | 0.5 mL |
| $V_3$ | 1.6 mL |

## References

1. S. E. Park, Epidemiology, virology, and clinical features of severe acute respiratory syndrome -coronavirus-2 (SARS-CoV-2; Coronavirus Disease-19). Clin Exp Pediatr, (2020).

2. D. Wang et al., Clinical Characteristics of 138 Hospitalized Patients With 2019 Novel Coronavirus-Infected Pneumonia in Wuhan, China. JAMA, (2020).

3. X. Yang et al., Clinical course and outcomes of critically ill patients with SARS-CoV-2 pneumonia in Wuhan, China: a single-centered, retrospective, observational study. Lancet Respir Med, (2020).

4. G. Onder, G. Rezza, S. Brusaferro, Case-Fatality Rate and Characteristics of Patients Dying in Relation to COVID-19 in Italy. JAMA, (2020).

5. K. S. o. I. D. a. K. C. f. D. C. a. Prevention, Analysis on 54 Mortality Cases of Coronavirus Disease 2019 in the Republic of Korea from January 19 to March 10, 2020. J Korean Med Sci 35(12), (2020).

6. C. W. Patrick GT Walker, Oliver Watson et al. “The Global Impact of COVID-19 and Strategies for Mitigation and Suppression. WHO Collaborating Centre for Infectious Disease Modelling, MRC Centre for Global Infectious Disease Analysis,” (Abdul Latif Jameel Institute for Disease and Emergency Analytics, Imperial College London, 2020).

7. B. E. Young et al., Epidemiologic Features and Clinical Course of Patients Infected With SARS-CoV-2 in Singapore. JAMA, (2020).

8. R. Wölfel et al., Virological assessment of hospitalized patients with COVID-2019. Nature, (2020).

9. J. Y. Kim et al., Viral Load Kinetics of SARS-CoV-2 Infection in First Two Patients in Korea. J Korean Med Sci 35, e86 (2020).

10. F. X. Lescure et al., Clinical and virological data of the first cases of COVID-19 in Europe: a case series. Lancet Infect Dis, (2020).

11. D. D. Ho et al., Rapid turnover of plasma virions and CD4 lymphocytes in HIV-1 infection. Nature 373, 123–126 (1995).

12. A. S. Perelson, A. U. Neumann, M. Markowitz, J. M. Leonard, D. D. Ho, HIV-1 dynamics in vivo: virion clearance rate, infected cell life-span, and viral generation time. Science 271, 1582–1586 (1996).

13. S. E. Holte, A. J. Melvin, J. I. Mullins, N. H. Tobin, L. M. Frenkel, Density-dependent decay in HIV-1 dynamics. J Acquir Immune Defic Syndr 41, 266–276 (2006).

14. A. P. Smith, D. J. Moquin, V. Bernhauerova, A. M. Smith, Influenza Virus Infection Model With Density Dependence Supports Biphasic Viral Decay. Front Microbiol 9, 1554 (2018).

15. B. Cao et al., A Trial of Lopinavir-Ritonavir in Adults Hospitalized with Severe Covid-19. N Engl J Med, (2020).

16. M. Wang et al., Remdesivir and chloroquine effectively inhibit the recently emerged novel coronavirus (2019-nCoV) in vitro. Cell Res 30, 269–271 (2020).

17. T. K. Warren et al., Therapeutic efficacy of the small molecule GS-5734 against Ebola virus in rhesus monkeys. Nature 531, 381–385 (2016).

18. J. T. Schiffer, D. A. Swan, L. Corey, A. Wald, Rapid viral expansion and short drug half-life explain the incomplete effectiveness of current herpes simplex virus 2-directed antiviral agents. Antimicrob Agents Chemother 57, 5820–5829 (2013).

19. J. T. Schiffer et al., Mathematical modeling of herpes simplex virus-2 suppression with pritelivir predicts trial outcomes. Sci Transl Med 8, 324ra315 (2016).

20. J. Chen et al., Cellular immune responses to severe acute respiratory syndrome coronavirus (SARS-CoV) infection in senescent BALB/c mice: CD4+ T cells are important in control of SARS-CoV infection. J Virol 84, 1289–1301 (2010).

21. R. Sanjuán, M. R. Nebot, N. Chirico, L. M. Mansky, R. Belshaw, Viral mutation rates. J Virol 84, 9733–9748 (2010).

22. M. L. Agostini et al., Coronavirus Susceptibility to the Antiviral Remdesivir (GS-5734) Is Mediated by the Viral Polymerase and the Proofreading Exoribonuclease. mBio 9, (2018).

23. D. B. Reeves et al., Mathematical modeling to reveal breakthrough mechanisms in the HIV Antibody Mediated Prevention (AMP) trials. PLoS Comput Biol 16, e1007626 (2020).

24. J. B. Antonio Gonçalves, Ruian Ke, Emmanuelle Comets, Xavier de Lamballerie, Denis Malvy, Andrés Pizzorno, Olivier Terrier, Manuel Rosa Calatrava, France Mentré, Patrick Smith, Alan S Perelson, Jérémie Guedj, Timing of antiviral treatment initiation is critical to reduce SARS-Cov-2 viral load. medRxiv 2020.04.04.20047886, (2020).

25. K. E. Kwang Su Kim, Yusuke Ito, Shoya Iwanami, Hirofumi Ohashi, Yoshiki Koizumi, Yusuke Asai, Shinji Nakaoka, Koichi Watashi, Robin N Thompson, Shingo Iwami, Modelling SARS-CoV-2 Dynamics: Implications for Therapy. medRxiv 2020.03.23.20040493, (2020).

26. J. D. Lundgren et al., Initiation of Antiretroviral Therapy in Early Asymptomatic HIV Infection. N Engl J Med 373, 795–807 (2015).

27. F. G. Hayden et al., Baloxavir Marboxil for Uncomplicated Influenza in Adults and Adolescents. N Engl J Med 379, 913–923 (2018).

28. S. Mulangu et al., A Randomized, Controlled Trial of Ebola Virus Disease Therapeutics. N Engl J Med 381, 2293–2303 (2019).

29. Z. Du et al., Serial Interval of COVID-19 among Publicly Reported Confirmed Cases. Emerg Infect Dis 26, (2020).

30. Y. Liu et al., Viral dynamics in mild and severe cases of COVID-19. Lancet Infect Dis, (2020).

31. O. Mitjà, B. Clotet, Use of antiviral drugs to reduce COVID-19 transmission. Lancet Glob Health, (2020).

32. K. S. Xue et al., Parallel evolution of influenza across multiple spatiotemporal scales. Elife 6, (2017).

33. L. Zou et al., SARS-CoV-2 Viral Load in Upper Respiratory Specimens of Infected Patients. N Engl J Med 382, 1177–1179 (2020).

34. K. P. Collins, K. M. Jackson, D. L. Gustafson, Hydroxychloroquine: A Physiologically-Based Pharmacokinetic Model in the Context of Cancer-Related Autophagy Modulation. J Pharmacol Exp Ther 365, 447–459 (2018).

35. R. J. De Boer, A. S. Perelson, Quantifying T lymphocyte turnover. J Theor Biol 327, 45–87 (2013).

36. A. S. Perelson et al., Decay characteristics of HIV-1-infected compartments during combination therapy. Nature 387, 188–191 (1997).

37. J. T. Schiffer et al., Mucosal host immune response predicts the severity and duration of herpes simplex virus-2 genital tract shedding episodes. Proc Natl Acad Sci U S A 107, 18973–18978 (2010).

38. P. L. Chan, P. Jacqmin, M. Lavielle, L. McFadyen, B. Weatherley, The use of the SAEM algorithm in MONOLIX software for estimation of population pharmacokinetic-pharmacodynamic-viral dynamics parameters of maraviroc in asymptomatic HIV subjects. J Pharmacokinet Pharmacodyn 38, 41–61 (2011).

39. M. Karlsson et al., Nonlinear mixed-effects modelling for single cell estimation: when, why, and how to use it. BMC Syst Biol 9, 52 (2015).

40. M. Lavielle, F. Mentré, Estimation of population pharmacokinetic parameters of saquinavir in HIV patients with the MONOLIX software. J Pharmacokinet Pharmacodyn 34, 229–249 (2007).

41. N. van Doremalen et al., Aerosol and Surface Stability of SARS-CoV-2 as Compared with SARS-CoV-1. N Engl J Med, (2020).

42. Thevarajan et al., Breadth of concomitant immune responses prior to patient recovery: a case report of non-severe COVID-19. Nature Medicine, (2020).

43. H. Akaike, in Second International Symposium on Information Theory, F. C. B.N. Petrov, Ed. (Akademiai Kiado, Budapest, 1973), pp. 267–281.

44. M. Frisk-Holmberg, Y. Bergqvist, E. Termond, B. Domeij-Nyberg, The single dose kinetics of chloroquine and its major metabolite desethylchloroquine in healthy subjects. Eur J Clin Pharmacol 26, 521–530 (1984).

45. Y. Huang et al., Population pharmacokinetics analysis of VRC01, an HIV-1 broadly neutralizing monoclonal antibody, in healthy adults. MAbs 9, 792–800 (2017).

46. J. Liu et al., Hydroxychloroquine, a less toxic derivative of chloroquine, is effective in inhibiting SARS-CoV-2 infection in vitro. Cell Discov 6, 16 (2020).

47. X. Yao et al., In Vitro Antiviral Activity and Projection of Optimized Dosing Design of Hydroxychloroquine for the Treatment of Severe Acute Respiratory Syndrome Coronavirus 2 (SARS-CoV-2). Clin Infect Dis, (2020).

